# Semantic Functioning in Temporal Lobe Epilepsy: A Systematic Review and Meta-Analysis

**DOI:** 10.1101/2025.09.17.25335967

**Authors:** Jacquie Eyres, Katia Lepore, Rubina Alpitsis, Terence J. O’Brien, Andrew Neal, Charles B. Malpas, Genevieve Rayner

## Abstract

**Background:** Despite the central role of the temporal lobes in conceptual processing, the impact of temporal lobe epilepsy (TLE) on semantic knowledge remains unclear.

**Objectives:** This systematic review and meta-analysis aimed to investigate semantic functioning in TLE and the impact that seizure lateralisation and surgical intervention have on semantic outcomes.

**Methodology:** A comprehensive literature search was conducted using Medline, Embase, and PsychINFO. Studies were eligible if they included participants aged ≥18 years with TLE and evaluated semantic functioning. Risk of bias was assessed using the Newcastle-Ottawa scale, and a narrative synthesis summarised the review findings. Meta-analyses compared semantic performance across individuals with TLE and healthy controls and left and right TLE.

**Results:** 141 studies, encompassing 8,241 participants (TLE: *n* = 5,623, controls: *n* = 2,618), were included, reporting over 30 different semantic measures. Both narrative review and meta-analysis showed significantly poorer semantic performance in people with TLE compared to controls, with impairments in semantic fluency (*g* = -1.35), WAIS-IV Vocabulary (*g* = -1.08), and Camels and Cactus Test (*g* = -1.37), but not on Pyramids and Palm Trees (*g* = -0.44). Some lateralisation effects were evident, with verbal semantic impairments more prominent in left TLE.

**Conclusions:** TLE is associated with a mild semantic impairment. While left-sided lesions are associated with worse verbal semantic impairment, lateralisation effects more broadly were mild and inconsistent. Our findings emphasise the need to conduct routine semantic assessments in TLE to support more precise cognitive deficit monitoring, better-informed surgical risk discussions, and the development of personalised rehabilitation plans.

**Highlights:** TLE is commonly associated with a mild impairment in semantic functioning.

LTLE is generally associated with relatively poorer performance on verbal-based semantic measures.

However, both LTLE and RTLE tend to demonstrate impairment across both verbal- and visual-based semantic measures.

In sum, semantic measures should be included in routine cognitive evaluations to support the clinical care of individuals with TLE.

## 1. Introduction

Temporal lobe epilepsy (TLE) is the most common form of focal epilepsy and is commonly associated with memory impairment.^1^ While episodic memory dysfunction is well-established in this population,^2^ there is a growing body of evidence suggesting that semantic memory, including the knowledge of facts, concepts, and word meaning, may also be compromised.^1^ The leading framework for semantic processing is the ‘hub-and-spoke’ model, ^3, 4^ which proposes that semantic knowledge is supported by a distributed neural network. The bilateral anterior temporal lobes serve as a central network ‘hub’, ^5, 6^ integrating modality-specific information from sensory, motor, and limbic cortices (i.e., the ‘spokes’) to form cohesive semantic representations.^7^ The current understanding of the role of the anterior temporal lobe in semantic processing is derived from conditions involving bilateral anterior temporal lobe damage (e.g., semantic dementia or herpes simplex encephalitis), where profound semantic loss is a central feature.^8–10^ Recent extensions of this model propose a degree of specialisation within and between the anterior temporal lobes due to asymmetric white matter connectivity.^11, 12^ For instance, the typically left-dominant hemisphere specialises in verbal semantics due to enhanced connectivity with language networks.^3, 4, 13^ Given that most prior work has focused on bilateral damage, any individual contributions of the left and right anterior temporal lobes remain unclear.^8–10^

The typically unilateral pathology of TLE offers a uniquely suited disease model for investigating how conceptual knowledge is organised within this bilateral network.^12^ Given the hypothesised role of the anterior temporal lobes in semantic processing, it follows that dysfunction in this region could significantly disrupt semantic function, whether due to the underlying epileptic network, epileptogenic lesions, effects of recurrent focal seizures, or surgical resection or ablation.

Yet, clinical investigations present a more complex picture. Intriguingly, patients with TLE rarely subjectively report semantic difficulties, more often describing episodic memory or naming issues.^1, 14^ Although some studies suggest intact semantic functioning in TLE,^15, 16^ the broader literature, particularly those with robust methodology and large samples, do suggest a mild semantic deficit.^12, 17, 18^ The relatively mild nature of these impairments compared to those observed in bilateral conditions, has led to the theory that the anterior temporal lobes function as undifferentiated, partially redundant ‘demi-hubs’ that are semi-robust to damage: unilateral injury results in modest impairment due to compensatory mechanisms of the contralateral hemisphere, whereas bilateral damage results in profound impairment due to a greater degree of network dysfunction.^7, 19^

This systematic review and meta-analysis investigates the presence of semantic impairment in TLE. Greater awareness of these deficits will provide valuable insights to enhance the clinical management of people with epilepsy, and, more broadly, may elucidate how conceptual knowledge is organised unilaterally within this bilateral system. The primary objectives were to evaluate semantic functioning across three key comparisons: i) individuals with TLE and healthy controls, ii) different lateralisation of TLE, and iii) pre- versus post-temporal lobe resection. A meta-analysis of studies reporting on several of the most commonly used semantic measures was also conducted.

## 2. Methodology

### 2.1 Registration and Reporting Guidelines

The research methodology was registered on PROSPERO on 25 June 2024 prior to data collection (registration number: CRD42024553990) and written in accordance with the Preferred Reporting Items for Systematic Review and Meta-Analysis (PRISMA) guidelines. As this review is based on secondary use of deidentified published data, ethical clearance was not required.

### 2.2 Eligibility Criteria

Studies were required to meet the following eligibility criteria: (1) the patient group must be diagnosed with TLE; (2) aged 18 years or older; (3) inclusion of at least one psychometric measure of semantic function; (4) reporting original data; and (5) published in English. The outcome measure was defined as any psychometric measure that assessed semantic functioning, regardless of standardisation or validity. Studies were excluded if (1) they involved non-human participants, or (2) participants had a concurrent neurological disorder with known cognitive symptomatology. No restrictions were placed on study design, sample size, patient demographics, or publication date. Only peer-reviewed, full-text articles were included.

### 2.3 Information Sources and Search Strategy

The search terms were formulated following consultation with a librarian specialising in systematic reviews. Relevant studies were identified through a systematic literature search of Medline, Embase, and PsychINFO electronic databases. The search strategy was as follows: (((“semantic fluency”) OR (“conceptual knowledge”) OR (“abstract concepts”) OR (“category fluency”) OR (“Controlled Oral Word Association Test”) OR (“COWAT”) OR (“Camels and Cactus”) OR (“Pyramids and Palm Trees”)) AND ((“epilepsy”) OR (“temporal lobe epilepsy”) OR (“TLE”) OR (“focal epilepsy”) OR (“epilepsy surgery”) OR (“anterior temporal lobectomy”) OR (“ATL”))). The period was from inception to 30th July 2025. Searches of grey literature, Google Scholar (for related articles), and article reference lists were conducted to identify additional articles not found in the primary searches.

### 2.4 Study Selection and Data Collection

Covidence systematic review software^20^ was used for data screening and extraction. After removing duplicate records, two reviewers (J.E. and K.L.) independently screened search results for eligibility in two stages: (1) title and abstract screening; and (2) full-text screening. Any conflicts were resolved by a third reviewer (G.R.). Data for the meta-analysis was independently collected by the two reviewers using a standardised data collection form, with disagreements resolved by consensus. Primary outcomes focused on the rate and nature of semantic impairment in temporal lobe epilepsy as defined by study methodology. Secondary outcomes included a comparison of performance between left and right TLE and pre- and post-surgery groups.

### 2.5 Study Quality Assessment

The reviewers also independently assessed study quality using an adapted Newcastle-Ottawa Quality Assessment Scale (NOS),^21^ a validated tool for evaluating bias in nonrandomised, observational studies. Studies were assessed on (A) participant selection, (B) study group comparability, (C) outcome assessment. A risk of bias score was assessed using a ‘star system’ for each category, where the greater the number of stars equates to a lower levels of bias. Studies were awarded stars for each applicable domain. Studies without a control group or longitudinal design had adjusted total scores. Disagreements were again resolved through consensus.

### 2.6 Data-Synthesis

Variability in methodology and outcome reporting limited meta-analysis across all semantic tasks within the review. Meta-analysis summarised results for measures with comparable methodology assessed in ≥3 studies.^22^ For each comparison, standardised mean differences were computed as Hedges’ *g*. As some studies contributed multiple effect sizes, a random-effects multi- level model accounted for within-study dependencies. The model was fitted using restricted maximum likelihood (REML) estimation in the *metafor* package in R software,^23^ with study and nested effect sizes specified as random effects. I^2^ was calculated and effect size was reported with 95% confidence intervals. Separate moderator analyses (namely, pre- and post-resection) were also conducted.^24^

### 2.7 Data Availability

The data used in this manuscript is available at: https://osf.io/10.17605/OSF.IO/UEVM3.

## 3. Results

### 3.1 Study Selection

After duplicates were removed, 1,153 potentially relevant studies were identified through database and hand searches of reference lists. Of these, 764 were excluded through title and abstract screening. At full-text review, an additional 248 studies were excluded, resulting in a final sample of 141 studies (see Figure 1).

**Figure 1.**
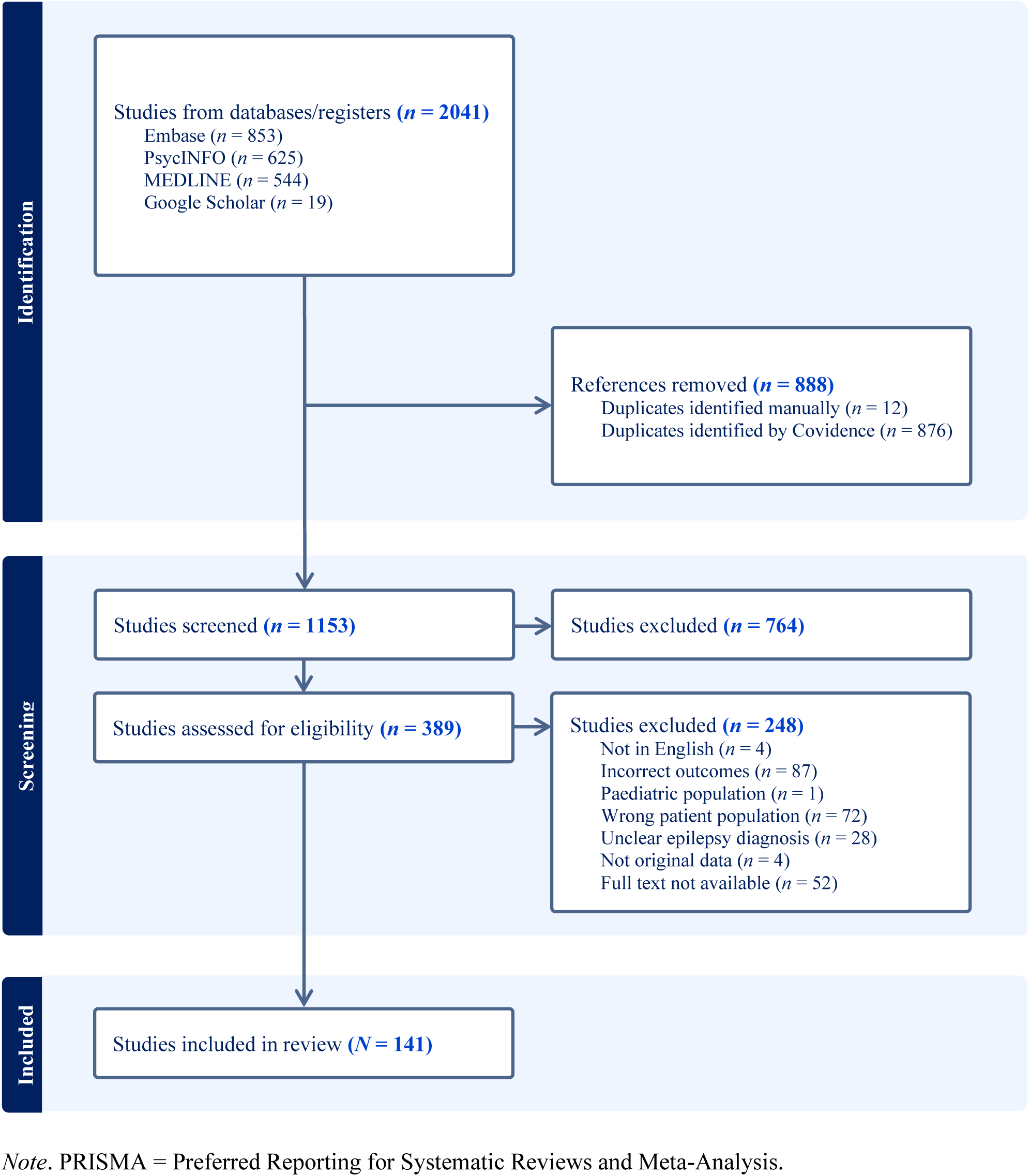
PRISMA Workflow of Literature Screening

### 3.2 Study Characteristics

Supplementary Table 1 summarises the included studies. A total of 141 studies, encompassing 8,241 participants (TLE *n* = 5,623, controls *n* = 2,618), were included. Sample size ranged from 1 to 415 across all participant groups (combined). Of the 411 studies, 16 had small total samples (*n* < 15), while the rest had medium or large samples (*n* ≥ 15). The mean age of TLE participants spanned 23.67 to 67.70 years, with a fairly balanced distribution of male and females. The mean age of controls ranged from 24.3 to 67.8 years, also with an even distribution of male and females. All studies included individuals with TLE (LTLE: *k* = 78, RTLE: *k* = 62) and 89 included healthy controls. Most participants were lesion-positive on MRI. Epilepsy onset ranged from 0.5 and 60.36 years, with a typical duration of 15-25 years. Mean IQ for the TLE groups ranged from 75.6 to 123.43 but was generally between 90-100. Education ranged from 6.93 to 17.10 years, but was most commonly high school or above. Of the included studies, 88 assessed pre-surgical patients, 16 post-surgical, and 25 compared pre- and post-surgery performance.

### 3.3 Semantic Functioning Outcome Measures

Over 30 different semantic measures were used across studies, spanning object knowledge, person knowledge, semantic association, priming, personal semantic knowledge, and general knowledge. The most common measure was semantic fluency from the Controlled Oral Word Association Test.^25^ However, within individual measures, there was variability across studies in task stimuli and instructions. For example, timings (e.g., 30, 60, 90, or 120 seconds for semantic fluency), stimuli (e.g., different celebrities in famous faces or different categories in fluency tasks), and the types of questions asked (e.g., names versus occupations in famous faces) differed within measures across studies.

### 3.4 Methodological Quality

Figure 2 and Supplementary Table 2 summarise risk of bias on a modified NOS scale. The total number of stars each study was eligible for varied according to study design. Most studies rated on this scale (136/141; 96%) were deemed as having a sufficiently representative sample. Where follow-up was present, the majority of studies employed a sufficiently long follow-up period for outcomes to occur, defined as ≥1 month after surgery (39/55; 71%). Additionally, most studies rated on this scale had sufficient follow-up rates, defined as 80% (18/24; 75%). Potential bias was most commonly introduced when studies failed to ensure comparability of the cohorts by controlling for differences in age, sex, or education.

**Figure 2.**
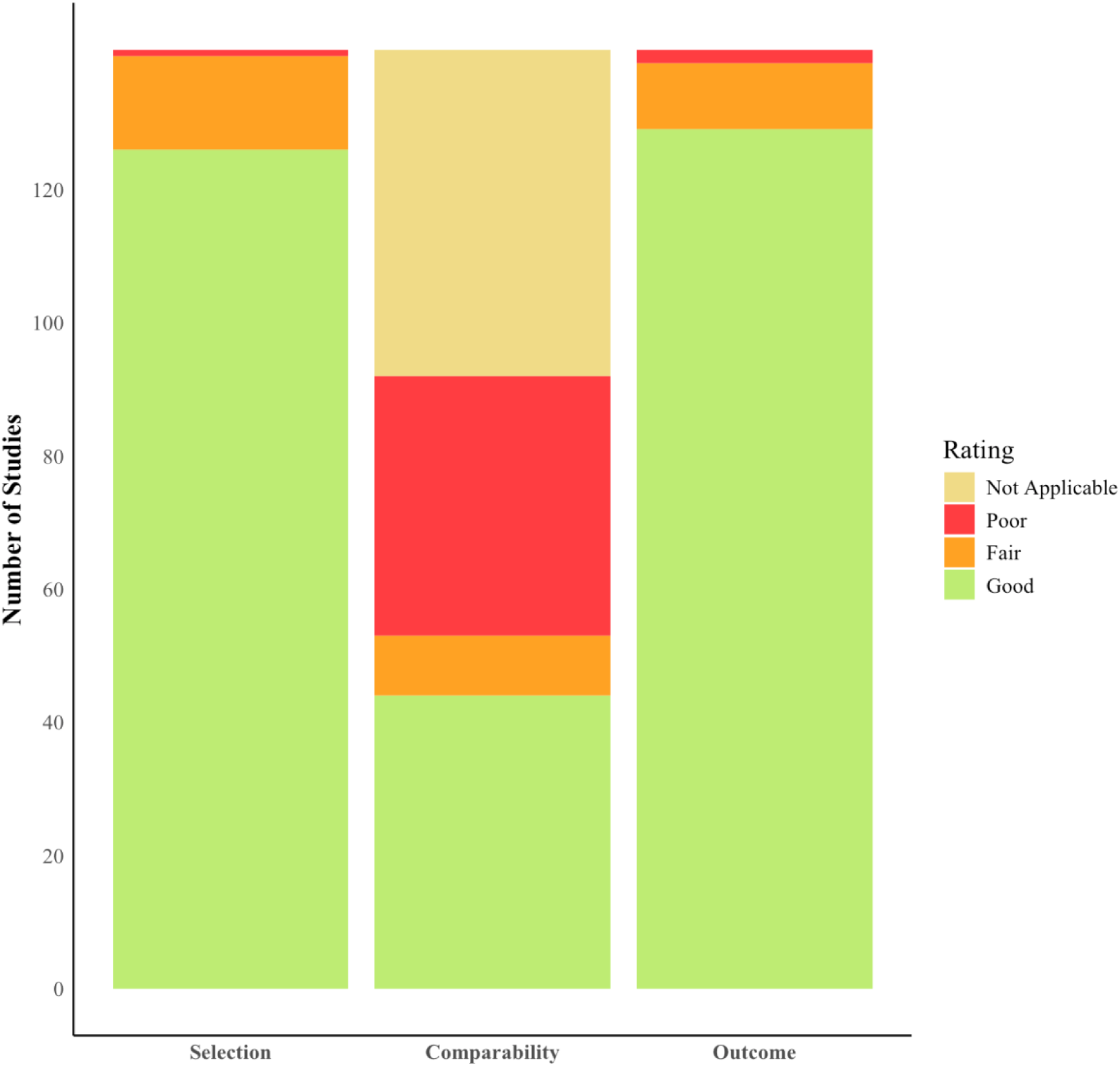
Study Quality Analysis Using the Newcastle-Ottawa Quality Assessment Scale

### 3.5 Review Findings

The following section summarises findings across various semantic task domains through narrative synthesis, followed by validation of these results via meta-analysis. A description summarising review findings is in Figure 3.

**Figure 3.**
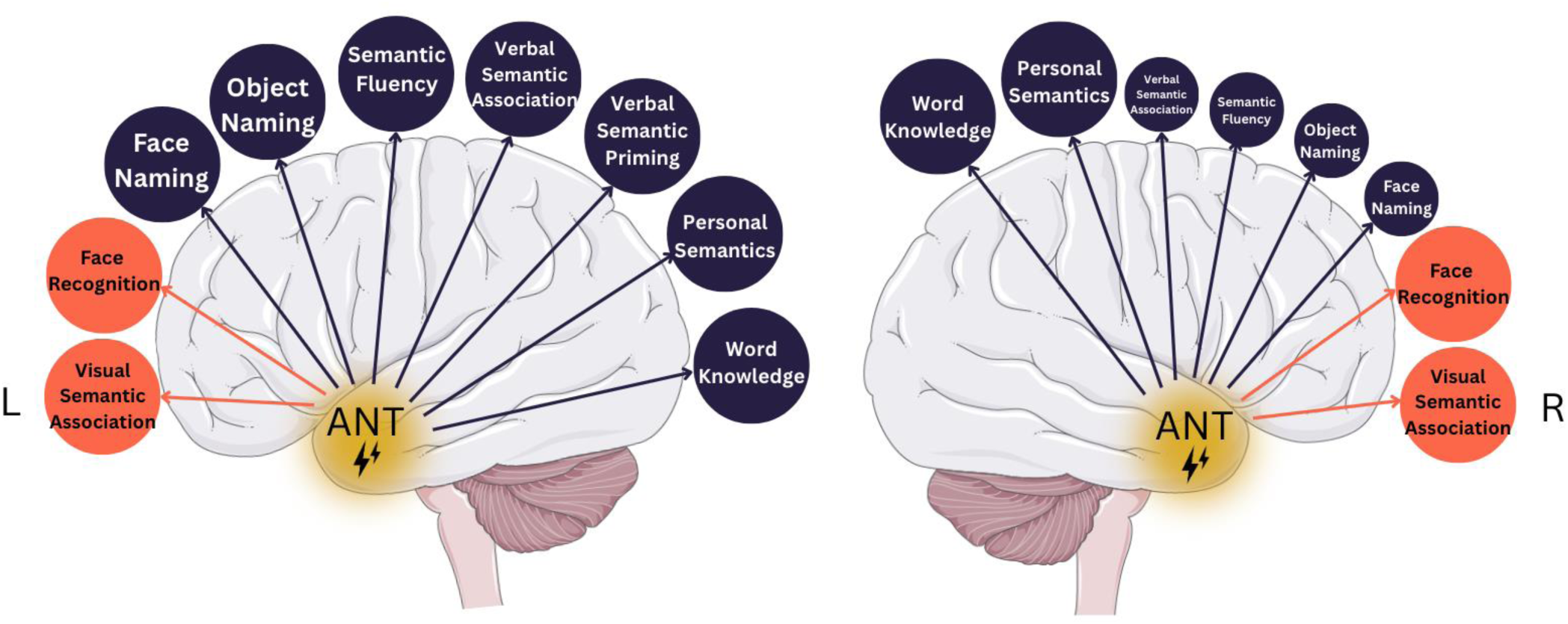
Lateralisation of Semantic Memory Function in Temporal Lobe Epilepsy (TLE): Summary of Empirical Evidence

#### 3.5.1 Narrative Synthesis

##### 3.5.1.1 Object Knowledge

Once other cognitive processes have been accounted for (e.g., visual-perception or phonological retrieval of words), poor naming performance has been interpreted as evidence of semantic dysfunction.^26^ TLE is commonly associated with naming impairments, particularly when the epileptogenic zone involves the posterior and basal temporal regions of the language-dominant hemisphere.^1,27^Although basic-level picture naming may be preserved in TLE,^13^ naming impairments are reliably identified on more sensitive measures designed to probe mild semantic loss,^12, 13, 18^ such as specific-level picture naming requiring subordinate-level responses (e.g., ‘poodle’ instead of ‘dog’). In our review, individuals with TLE routinely demonstrate poorer naming performance compared to healthy controls.^12, 13, 18, 28–30^ Consistent with established findings implicating the left hemisphere in language-based tasks,^11^ our review found strong evidence for poorer naming performance in LTLE compared to RTLE.^13, 24,31^ Additionally, naming performance is more routinely compromised in patients who have undergone temporal lobe resection^12, 13, 18, 28^ compared to those who are surgically-naïve.^29–34^ Together, these findings suggest that impaired semantic knowledge may contribute to the some of the naming difficulties observed in TLE, and particularly LTLE.

##### 3.5.1.2 Person Knowledge

Accurate face naming requires both facial recognition and name retrieval.^35^ Our review identified that, compared to controls, people with TLE consistently demonstrate poorer performance on the Famous Faces Task, ^12, 17, 36–44^ assessing the ability to recognise and name well-known individuals. The individual stages of famous face processing has been used to examine the hemispheric specialisation hypothesis, with familiarity judgements positively associated with grey-matter volume in the right anterior middle temporal gyrus, and person naming associated with left anterior temporal volume.^45^ In support of this theory, our review found that people with TLE, and particularly those with RTLE, demonstrate greater facial recognition impairments than healthy controls.^46–49^ Additionally, although some studies report no differences between TLE subgroups,^47, 49, 50^ most studies provide support for left-hemispheric dominance for person-specific naming, with greater impairments identified in LTLE.^12, 13, 44, 46, 48^ Collectively, studies within our review provide some evidence for a processing bias for person knowledge, with the right temporal lobe demonstrating greater involvement in facial recognition and the left temporal lobe in person name retrieval.

##### 3.5.1.3 Category Fluency

Semantic fluency tasks, in which individuals generate as many words as possible from a given category within a time limit, are one the most widely used semantic measures.^25^ Across studies, there is overwhelming evidence that people with TLE exhibit poorer semantic fluency performance than healthy controls,^31, 48, 51–56^ consistently producing fewer exemplars across a broad range of categories (e.g., animals, fruits, cars, living things, non-living things) both pre- and post-resection. The small number of studies reporting comparable performance with controls, ^34, 40, 57–59^ were often limited by small samples or did observe a non-significant trend for poorer TLE performance. With respect to lateralisation, LTLE routinely demonstrate greater semantic fluency impairment than RTLE, ^60–64^ supporting the proposed role of the left temporal lobe in verbal semantic processing. Studies directly comparing pre- and post-surgical semantic fluency performance are mixed. Despite comparable sample sizes and methodologies, post-operative outcomes vary, with some studies reporting stable performance,^40, 65–67^ while others show either improvement^57, 68, 69^ or decline.^70^ Interestingly, some studies suggest an initial trend towards non-significant post-operative improvement,^67, 71–73^ which becomes significant at longer follow-up intervals.^31, 74–76^ Overall, our review identified clear evidence for semantic fluency impairments in TLE, with LTLE showing greater deficits than RTLE. While post-surgical outcomes are mixed, many studies suggest a trend towards improvement over time.

##### 3.5.1.4 Semantic Association

The Pyramid and Palm Trees Test (PPT),^77^ and Camel and Cactus Test (CCT)^78^ are commonly used to assess visual semantic association in TLE. Both tasks require participants to select the picture most semantically related to a target from multiple-choice options. Throughout our review, performance on the PPT was mixed; while many smaller studies report comparable performance between TLE and controls,^36, 41, 79^ larger studies identified significant impairments.^28, 33, 80^ In contrast, people with TLE consistently perform worse than controls on the CCT,^62, 81–83^ suggesting that the CCT, offering four response options (chance level = 0.25), may be a more sensitive measure of verbal semantic association than the PPT, which has only two response options (chance level = 0.5). Interestingly, one study comparing LTLE and RTLE on visual semantic association, found individuals with RTLE outperformed those with LTLE.^84^ Individuals with TLE also exhibit poorer performance on verbal semantic association tasks (e.g., the Synonym Judgment Task) in accuracy and speed compared to healthy controls.^12, 13, 17, 18^ When examining TLE subgroups, people with LTLE typically demonstrate worse performance than controls,^12, 13, 17, 18^ whereas RTLE performance was more variable. One study reported RTLE deficits,^12^ whereas others with comparable samples found no significant differences.^12, 18^ Concordant with outcomes observed in other verbal-based semantic tasks, LTLE generally perform worse than RTLE on verbal semantic association measures.^12, 85^ Together, these results suggest that visual and verbal semantic association impairments are common in TLE, and that LTLE may be more susceptible to deficits in verbal semantic association.

##### 3.5.1.5 Priming

Network-based models of semantic functioning^86^ propose that conceptual knowledge is accessed through the spreading activation of connected nodes within the semantic network. The functionality of this network is commonly probed using semantic priming paradigms, where the presentation of semantically-related prime words generates faster response times to target words. Early work from Blaxton et al. (1993)^87^ and Miyamoto et al. (1995)^88^ demonstrated slower reactions to primed targets in individuals with TLE compared to healthy controls, indicating a compromised semantic network. The single study comparing priming effects in LTLE and RTLE performance found no significant differences in reaction time or accuracy between TLE subgroups.^89^ Despite limited research investigating semantic priming in TLE, the existing literature suggests that temporal lobe dysfunction may be associated with disruptions to the knowledge store or the activation process within the semantic network.

##### 3.5.1.6 Personal Semantic Knowledge

Factual knowledge about oneself (e.g., birth date, address), may also be affected by temporal lobe damage.^41^ In our review, evidence regarding personal semantic performance in TLE is mixed. Among the 13 studies comparing autobiographical semantic knowledge in TLE to controls, seven reported comparable performance^40–42, 44, 79, 90–92^ and six identified poorer performance.^34, 48, 93–95^ Also, when studies assessed personal semantics across periods of the lifespan, there were inconsistent findings with regards to whether childhood, early adulthood, or recent autobiographical knowledge was impaired.^40, 41, 65, 94, 95^ When comparing TLE subgroups, the majority of studies failed to find a difference between LTLE and RTLE on autobiographical semantic knowledge.^44, 90–93, 95^ Despite mixed findings, this review identified some evidence that TLE may affect personal semantic knowledge.

##### 3.5.1.7 General Knowledge

Temporal lobe dysfunction may also be associated with impaired general knowledge (e.g., knowing that the Earth has seven continents), with Blaxton (1992)^96^ and Delazer et al. (2004)^97^ both finding that individuals with TLE had poorer factual knowledge than healthy controls. Beyond just naming issues, TLE may also be associated with reduced knowledge of word meaning, frequently producing vague definitions when defining common words.^28, 29, 56, 89, 98–100^ Intriguingly, people with LTLE and RTLE often perform similarly on word definition tasks, even after resective surgery.^101, 102^ Together, these findings suggest that temporal lobe dysfunction in either hemisphere may contribute to reduced factual knowledge.

#### 3.5.2 Meta-Analysis

Meta-analytic results for semantic performance in TLE across semantic measures are summarised in Table 1.

**Table 1.**
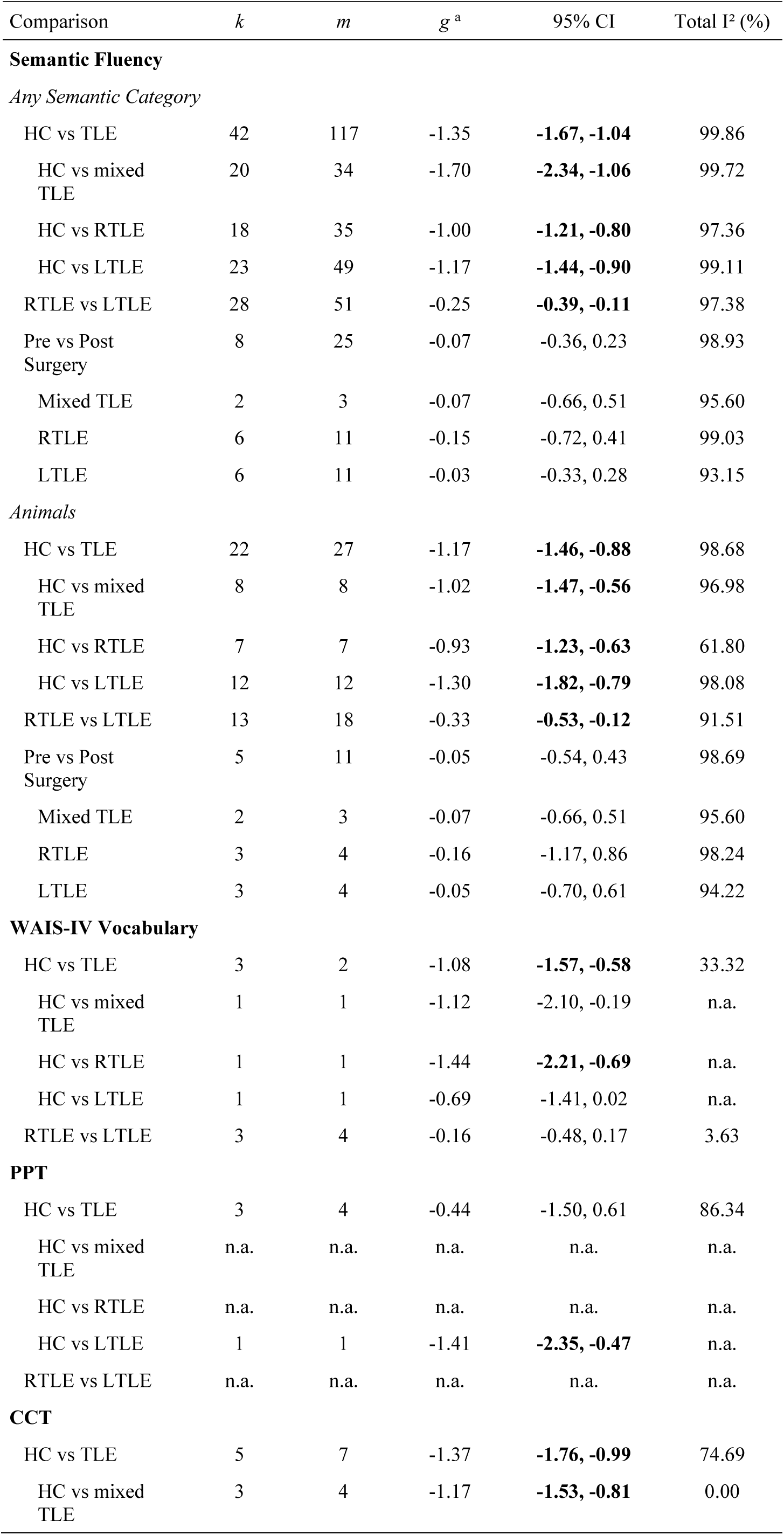
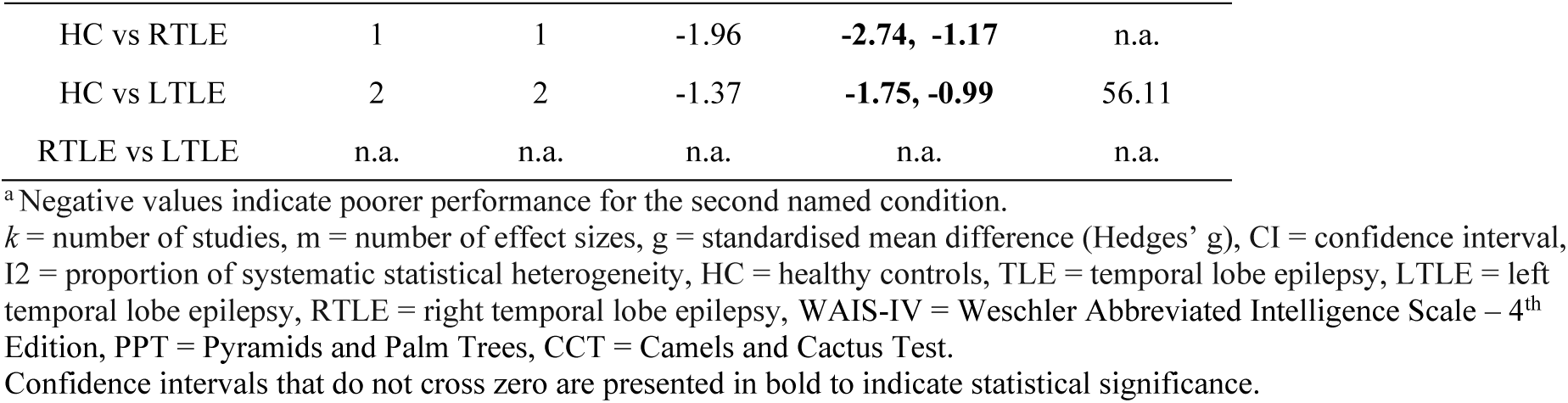
Summary results of main and sub-group analyses.

##### 3.5.2.1 Category Fluency

Commensurate with narrative review findings, patients with TLE performed significantly worse than healthy controls across semantic fluency measures (*g* = -1.35, 95% CI [-1.67, -1.04]). In particular, mixed TLE (*g* = -1.70, 95% CI [-2.34, -1.06]), LTLE (*g* = -1.17, 95% CI [-1.44, -0.90]), and RTLE groups (*g* = 1.00, 95% CI [-1.21, -0.80]) performed significantly worse than controls. With respect to lateralisation, LTLE performed significantly worse than RTLE (*g* = -0.25, 95% CI [-0.39, - 0.11]). No significant differences were observed pre- versus post-surgery across subgroups (all *p* > .05). A test of moderators, including surgery status and semantic category (i.e., living, non-living, mixed), did not significantly explain additional variability in semantic fluency performance between TLE and controls and between LTLE and RTLE (both *p* > .05). A similar pattern of results was observed for animal fluency, the most common category used in semantic fluency.

##### 3.5.2.2 General Knowledge

Similar to conclusions drawn from narrative synthesis, patients with TLE performed significantly worse than controls (*g* = -1.08, 95% CI [-1.57, -0.58]) on WAIS-IV Vocabulary. RTLE differed significantly from controls (*g* = -1.44, 95% CI [-2.21, -0.69]), while LTLE did not (*g* = -0.69, 95% CI [-1.41, 0.02]). With respect to lateralisation, there was no significant difference in performance between LTLE and RTLE (*g* = -0.16, 95% CI [-0.48, 0.17]). A test of moderators, including surgery status, did not significantly explain additional variability in Vocabulary performance between LTLE and RTLE (*p* > .05). Insufficient studies were available to examine pre- versus post-surgery Vocabulary performance.

##### 3.5.2.3 Visual Semantic Association

While patients with TLE performed significantly worse than controls on the CCT (*g* = -1.37, 95% CI [-1.76, -0.99]), no significant difference were observed on the PPT (*g* = -0.44, 95% CI [-1.50, 0.61]), consistent with narrative review findings. Surgery status did not significantly explain additional variability in CCT (*p* = 0.120) performance between TLE and controls (*p* > .05), but did for PPT (*p* = .029), with poorer outcomes in the post-surgical group than the mixed group (containing both pre- and post-surgical cases). No significant difference emerged between the pre- and post-surgical groups (*p* > .05). Insufficient studies were available to examine LTLE versus RTLE and pre- versus post-surgery PPT or CCT performance.

## 4. Discussion

This systematic review and meta-analysis examined semantic functioning in TLE. We examined data from 141 studies assessing semantic functioning in 5,623 individuals diagnosed with TLE. Despite our sample size suggesting a relatively robust body of research, there is a shortage of targeted investigations into semantic functioning in TLE, with many of the data in this review arising from a small number of semantic measures included in broader cognitive test batteries. Nonetheless, the available evidence provides overwhelming support for the presence of mild semantic impairments in TLE. This review provides some evidence for the presence of lateralisation effects, with LTLE associated with relatively poorer performance on verbal-based semantic measures, and RTLE commonly exhibiting difficulties with facial recognition. However, the strength of these lateralisation effects was mild, and mixed results were not uncommon. Similarly, our understanding of the impact of surgical treatment for TLE on semantic function is equivocal, with some studies reporting post- operative improvement, while others suggest stable performance, or even decline. The following section discusses the need to broaden our understanding of memory impairment in TLE to include semantic dysfunction, followed by an exploration of the evidence for lateralisation effects, possible implications of our findings, and directions for future research.

### 4.1 Broadening the Memory Phenotype in TLE to Include Semantic Dysfunction

The central finding of this review is that TLE is associated with impairments in semantic functioning, consistently underperforming relative to controls across a broad range of semantic measures. These findings reinforce the well-established notion that memory impairments are a common cognitive consequence of TLE,^1^ and suggest that the profile of also encompasses semantic deficits.^15, 16^ Given that many semantic tasks are multidetermined, requiring both stored knowledge and executive-based retrieval processes, poor semantic performance may not be solely attributable to temporal lobe dysfunction.^103^ However, executive dysfunction alone is unlikely to account for the pattern of widespread impairment observed across a broad range of semantic tasks.^103^ This is supported by evidence that tasks with minimal executive demands (e.g., word-picture matching) are usually preserved in individuals with frontal lobe pathology.^26^ Even in tasks with greater executive demands (e.g., semantic fluency), comparisons between individuals with frontal and temporal lobe damage consistently show that intact performance relies more heavily on semantic knowledge systems supported by the temporal lobes rather than frontally-mediated strategic-search functions.^104–106^ Overall, our findings provide support for the role of the anterior temporal lobe in semantic processing,^16^ highlighting the importance of including semantic measures in neuropsychological evaluations of temporal lobe pathology.

### 4.2 Inconsistent Laterality Effect in TLE

Although our review provides some evidence for a hemispheric processing bias,^4, 13^ our findings also suggest overlapping functions within the left and right anterior temporal lobes.^15^ Consistent with findings observed in semantic dementia cases with greater left-sided atrophy,^78^ we found that people with LTLE generally exhibit more pronounced deficits on verbally-mediated semantic tasks (e.g., object or person naming, category fluency, synonym judgement). While RTLE is associated with relatively greater difficulty with facial recognition, we did not consistently observe disproportionately poorer RTLE performance on other visually-mediated semantic measures (e.g., visual association). Although the functions of the anterior temporal lobes have been hypothesised to be largely comparable between the hemispheres, Lambon-Ralph et al. (2010) propose that differences in white-matter connections between the left and right anterior temporal lobes and broader modality-specific brain regions may explain the presence of subtle modality-specific processing biases following unilateral damage. For instance, due to stronger connections with typically left-lateralised language networks, left anterior temporal lobe pathology would be associated poorer performance on language-based semantic tasks. ^3, 4, 13^ Also consistent with our findings, right-sided lesions have been proposed to be associated with comparatively poorer performance on facial recognition tasks due to relatively greater connections between the fusiform face area in the right temporal lobe.^107^

Despite some evidence in support of hemispheric processing biases, both patient groups demonstrated widespread impairment across semantic measures, irrespective of input modality. The hypothesis that the anterior temporal lobes are redundant is partially challenged by this finding,^7^ as the intact lobe does not appear to fully compensate for the loss of function in the damaged lobe. Using language-based measures as an example, both LTLE and RTLE showed marked impairment on several verbal tasks (e.g., object naming, category fluency, vocabulary) compared to controls. Despite LTLE typically having relatively worse performance, the difference between TLE groups was relatively small. While epilepsy-related factors (e.g., anti-seizure medication or seizure burden) could contribute to poor RTLE performance on language-based tasks,^108^ these variables were generally comparable between TLE groups. Together, these findings suggest that impaired performance on verbal-based semantic tasks are not exclusive to individuals with TLE in the language-dominant hemisphere, raising questions about the lateralising utility of verbal semantic impairment in TLE.

Successful performance across many semantic measures likely requires bilateral activation, as different task demands may rely on separate modality-specific neural networks.^12, 26^ As an illustration, as the right temporal lobe is hypothesised to be critical for facial recognition,^109^ while the left temporal lobe plays a greater role in person name retrieval, double dissociations may be present on the Famous Faces task. ^11^ Drane et al. (2013)^46^ supports this, with LTLE associated with intact facial recognition but impaired naming performance, and RTLE demonstrating the opposite pattern. Recognition performance significantly improved in RTLE following lexical cueing, even for faces initially rated as unfamiliar, suggesting participants had difficulty processing visually presented information rather than a total loss of semantic knowledge. These findings highlight the importance of considering individual task demands when analysing the pattern of semantic impairments observed in TLE.

### 4.3 Mixed Post-Surgical Semantic Performance Outcomes

Historically, unilateral temporal lobe resections have been regarded as relatively benign in terms of semantic function, given that patients do not develop a ‘semantic dementia-like’ syndrome post-surgery.^15, 16^ Our findings present a more complex picture, with some patients demonstrating improved or stable post-operative performance and others experiencing mild reductions. Improved post-operative performance has been theorised to result from overall improvements in brain function resulting from reduced seizure activity.^110^ Relatively preserved performance following unilateral resection has been interpreted as evidence for redundancy of the anterior temporal lobes, whereby the intact lobe compensates for loss of function in the resected region.^7^ This redundancy has been hypothesised to produce a pattern of ‘graceful degradation’ in the face of damage, where minor unilateral pathology produces very subtle inefficiencies and larger lesions produce greater impairment, particularly if bilateral structures are compromised.^19^ However, unless the left and right anterior temporal lobes have completely identical functions, significant unilateral damage (e.g., large anterior temporal resection) would be expected to produce some degree of postoperative decline.^19^ Given the variability in post-surgical outcomes, further research investigating possible predictors of post-operative performance, including the extent of tissue resection, is needed.^111^

### 4.4 Clinical and Theoretical Implications

There are several clinical and theoretical implications of our findings. Given that TLE is associated with semantic impairment, our findings highlight the need for inclusion of semantic measures in routine neuropsychological evaluation. This will enable clinicians to more accurately assess for and monitor cognitive decrements associated with TLE, facilitate better-informed pre-surgical counselling regarding cognitive risk, and develop tailored prehabilitation and rehabilitation programs for semantic impairment. From a theoretical perspective, the presence of widespread impairment in left and right TLE for both verbal and visual information, supports the hypothesis that efficient semantic processing requires bilateral anterior temporal lobe involvement. ^5, 6^ ^11^ Despite this, the presence of relatively greater verbal semantic impairment in LTLE and facial recognition impairment in RTLE suggests that the functions of the bilateral anterior temporal lobes are not entirely identical.^11^ As such, our findings support the theory that subtle processing biases emerge within the anterior temporal lobes due to differences in white-matter connectivity to broader cognitive networks.^4, 13^

### 4.5 Review Limitations and Future Directions

Our study has several limitations that may be addressed in future work. Firstly, given the small number of studies comparing pre- and post-operative semantic performance in this review, future research should employ longitudinal designs to clarify the impact of surgical treatment on semantic functioning.^112^ Secondly, as most semantic measures were designed to identify semantic impairments in dementia cohorts, they may not be sensitive enough to detect the typically mild impairments observed in TLE. Future studies should adopt protocols used in early-stage semantic dementia research, which use more sensitive metrics (e.g., reaction times over accuracy) and measures (e.g., requiring more specific responses or presenting lower-frequency concepts) to detect subtle semantic deficits.^14, 113–115^ Thirdly, many studies in this review included clinically heterogenous samples, potentially biasing our pooled analyses. The diagnosis of TLE includes a broad range of subtypes with subtle differences in the affected cognitive networks (e.g., temporal pole, restricted mesial temporal, posterior temporal, basal temporal, anterior-mesial). Although most of our sample likely had unilateral anterior-mesial TLE given the high MRI-positive rate (likely higher than contemporary cohorts where MRI-negative cases are referred to stereo-EEG), subtypes were often not reported, and participants were routinely classified into a single broad TLE group. Additionally, many studies grouped LTLE and RTLE together, as well as surgically-naive and surgically-treated cohorts, limiting our understanding of lateralised differences or surgical consequences on semantic functioning. Moreover, as variability in surgical approaches confer different levels of cognitive risk, with selective amygdalohippocampectomy associated with better cognitive outcomes than the less precise anterior temporal lobectomy,^116^ future research should consider individual differences in TLE diagnoses and treatment approaches to help clarify possible predictors of semantic impairment.

Finally, as publication bias statistics are limited for data with multi-level dependencies, we cannot exclude the presence of publication bias in this review.

### 4.6 Conclusions

In conclusion, this review demonstrates that semantic impairment is a core feature of the cognitive profile of TLE. Our results provide evidence for bilateral anterior temporal lobe involvement in semantic processing, with damage to either the left or right temporal lobe associated with a mild impairment on both visual and verbal semantic tasks. Despite this, our results provide partial evidence for hemispheric processing biases, with TLE localised to the language-dominant left hemisphere demonstrating relatively greater impairment on verbal semantic tasks. The argument for aright-hemispheric bias towards visual semantics, however, is less clear. Our results suggest that the absence of a profound a ‘semantic dementia-like’ syndrome in unilateral TLE may be explained by (i) a more restricted area of network disruption compared to that observed in bilateral and neurodegenerative conditions, and (ii) a degree of functional compensation by the unaffected lobe.

Given an increased vulnerability for semantic impairment in unilateral TLE, our study highlights the importance of incorporating semantic measures in routine neuropsychological assessment batteries to support the clinical care and management of this cohort.

## Data Availability

The data used in this manuscript is available on Open Science Framework website.

https://osf.io/10.17605/OSF.IO/UEVM3.

## Acknowledgment

The authors thank librarian Vesna Birkic from The University of Melbourne for her advice on undertaking the search strategy for the systematic review.

## References

1. Helmstaedter C. Neuropsychological aspects of epilepsy surgery. Epilepsy & Behavior. 2004;5:45–55.

2. Giovagnoli AR, Erbetta A, Villani F, Avanzini G. Semantic memory in partial epilepsy: verbal and non-verbal deficits and neuroanatomical relationships.Neuropsychologia. 2005;43(10):1482–92. doi:10.1016/j.neuropsychologia.2004.12.010

3. Mion M, Patterson K, Acosta-Cabronero J, et al. What the left and right anterior fusiform gyri tell us about semantic memory. Brain. 2010;133(11):3256–3268.

4. Rice GE, Lambon Ralph MA, Hoffman P. The roles of left versus right anterior temporal lobes in conceptual knowledge: an ALE meta-analysis of 97 functional neuroimaging studies. Cerebral Cortex. 2015;25(11):4374–4391.

5. Ralph MAL, Jefferies E, Patterson K, Rogers TT. The neural and computational bases of semantic cognition. Nature reviews neuroscience. 2017;18(1):42–55.

6. Patterson K, Nestor PJ, Rogers TT. Where do you know what you know? The representation of semantic knowledge in the human brain. Nature reviews neuroscience. 2007;8(12):976–987.

7. Rogers TT, Lambon Ralph MA, Garrard P, et al. Structure and deterioration of semantic memory: a neuropsychological and computational investigation. Psychological review. 2004;111(1):205.

8. Binney RJ, Ralph MAL. Using a combination of fMRI and anterior temporal lobe rTMS to measure intrinsic and induced activation changes across the semantic cognition network. Neuropsychologia. 2015;76:170–181.

9. Jung J, Lambon Ralph MA. Mapping the dynamic network interactions underpinning cognition: a cTBS-fMRI study of the flexible adaptive neural system for semantics. Cerebral Cortex. 2016;26(8):3580–3590.

10. Noppeney U, Patterson K, Tyler LK, et al. Temporal lobe lesions and semantic impairment: a comparison of herpes simplex virus encephalitis and semantic dementia. Brain. 2007;130(4):1138–1147.

11. Rice GE, Hoffman P, Lambon Ralph MA. Graded specialization within and between the anterior temporal lobes. Annals of the New York Academy of Sciences. 2015;1359(1):84–97.

12. Rice GE, Caswell H, Moore P, Hoffman P, Lambon Ralph MA. The Roles of Left Versus Right Anterior Temporal Lobes in Semantic Memory: A Neuropsychological Comparison of Postsurgical Temporal Lobe Epilepsy Patients. Cerebral cortex (New York, NY : 1991). 2018;28(4):1487-1501. doi:10.1093/cercor/bhx362

13. Lambon Ralph MA, Ehsan S, Baker GA, Rogers TT. Semantic memory is impaired in patients with unilateral anterior temporal lobe resection for temporal lobe epilepsy. Brain : a journal of neurology. 2012;135(Pt 1):242–58. doi:10.1093/brain/awr325

14. Lambon Ralph MA, Ehsan S, Baker GA, Rogers TT. Semantic memory is impaired in patients with unilateral anterior temporal lobe resection for temporal lobe epilepsy. Brain. 2012;135(1):242–258.

15. Kho KH, Indefrey P, Hagoort P, Van Veelen C, van Rijen PC, Ramsey NF. Unimpaired sentence comprehension after anterior temporal cortex resection. Neuropsychologia. 2008;46(4):1170–1178.

16. Simmons WK, Martin A. The anterior temporal lobes and the functional architecture of semantic memory. Journal of the International Neuropsychological Society.2009;15(5):645–649.

17. Rouse MA, Ramanan S, Halai AD, et al. The impact of bilateral versus unilateral anterior temporal lobe damage on face recognition, person knowledge and semantic memory. medRxiv. 2024;((Rouse, Ramanan, Halai, Patterson, Rowe, Lambon Ralph) MRC Cognition and Brain Sciences Unit, University of Cambridge, Cambridge CB2 7EF, United Kingdom(Volfart) Universite de Lorraine, CNRS, Nancy F-54000, France(Volfart) Psychological Sciences Research)doi:10.1101/2024.02.10.24302526

18. Visser M, Forn C, Lambon Ralph MA, et al. Evidence for degraded low frequency verbal concepts in left resected temporal lobe epilepsy patients. Neuropsychologia. 2018;114(0020713, nzn):88-100.doi:10.1016/j.neuropsychologia.2018.04.020

19. Lambon Ralph MA, Cipolotti L, Manes F, Patterson K. Taking both sides: do unilateral anterior temporal lobe lesions disrupt semantic memory? Brain. 2010;133(11):3243–3255.

20. 20. *Covidence systematic review software*. Veritas Health Innovation; www.covidence.org

21. Wells GA, Shea B, O’Connell D, et al. The Newcastle-Ottawa Scale (NOS) for assessing the quality of nonrandomised studies in meta-analyses. 2000;

22. Ahn E, Kang H. Introduction to systematic review and meta-analysis. Korean journal of anesthesiology. 2018;71(2):103–112.

23. Viechtbauer W. Conducting meta-analyses in R with the metafor package. Journal of statistical software. 2010;36:1–48.

24. Higgins JP, Thompson SG, Deeks JJ, Altman DG. Measuring inconsistency in meta-analyses. bmj. 2003;327(7414):557-560.

25. Benton A, Hamsher dS, Sivan A. Controlled oral word association test. Archives of Clinical Neuropsychology. 1994;

26. Doughty O, Done D. Is semantic memory impaired in schizophrenia? A systematic review and meta-analysis of 91 studies. Cognitive Neuropsychiatry. 2009;14(6):473–509.

27. Trebuchon-Da Fonseca A, Guedj E, Alario FX, et al. Brain regions underlying word finding difficulties in temporal lobe epilepsy. Brain : a journal of neurology. 2009;132(Pt 10):2772–84. doi:10.1093/brain/awp083

28. Antonucci S, Beeson P, Labiner D, Rapcsak S. Lexical retrieval and semantic knowledge in patients with left inferior temporal lobe lesions. Aphasiology. 2008;22(3):281–304. doi:10.1080/02687030701294491

29. Caciagli L, Paquola C, He X, et al. Disorganization of language and working memory systems in frontal versus temporal lobe epilepsy. Brain : a journal of neurology. 2023;146(3):935–953. doi:10.1093/brain/awac150

30. Pope RA, Thompson PJ, Rantell K, Stretton J, Wright M-A, Foong J. Frontal lobe dysfunction as a predictor of depression and anxiety following temporal lobe epilepsy surgery. Epilepsy research. 2019;152(ema, 8703089):59-66. doi:10.1016/j.eplepsyres.2019.03.003

31. Sablik M, Fleury MN, Binding LP, et al. LonglJterm neuroplasticity in language networks after anterior temporal lobe resection. Epilepsia. 2025;66(1):207–225.

32. Giovagnoli AR. Characteristics of verbal semantic impairment in left hemisphere epilepsy. Neuropsychology. 2005;19(4):501–8. doi:10.1037/0894-4105.19.4.501

33. Giovagnoli AR, Villani F, Bell B, Erbetta A, Avanzini G. The chicken with four legs: a case of semantic amnesia and cryptogenic epilepsy. Epilepsy & behavior : E&B. 2009;14(1):261–8. doi:10.1016/j.yebeh.2008.09.015

34. Milton F, Muhlert N, Pindus DM, et al. Remote memory deficits in transient epileptic amnesia. Brain : a journal of neurology. 2010;133(Pt 5):1368–79.doi:10.1093/brain/awq055

35. Marful A, Paolieri D, Bajo MT. Is naming faces different from naming objects? Semantic interference in a face-and object-naming task. Memory & Cognition. 2014;42:525–537.

36. Condret-Santi V, Barragan-Jason G, Valton L, et al. Object and proper name retrieval in temporal lobe epilepsy: a study of difficulties and latencies. Epilepsy research. 2014;108(10):1825–38. doi:10.1016/j.eplepsyres.2014.09.001

37. Drane DL, Ojemann GA, Aylward E, et al. Category-specific naming and recognition deficits in temporal lobe epilepsy surgical patients. Neuropsychologia. 2008;46(5):1242–55. doi:10.1016/j.neuropsychologia.2007.11.034

38. Ellis AW, Young AW, Critchley EM. Loss of memory for people following temporal lobe damage. Brain : a journal of neurology. 1989;112 (Pt 6)(0372537, b5f):1469–83. doi:10.1093/brain/112.6.1469

39. Griffith HR, Richardson E, Pyzalski RW, et al. Memory for famous faces and the temporal pole: functional imaging findings in temporal lobe epilepsy. Epilepsy & behavior : E&B. 2006;9(1):173–80. doi:10.1016/j.yebeh.2006.04.024

40. Manning L, Denkova E, Unterberger L. Autobiographical significance in past and future public semantic memory: a case-study. Cortex; a journal devoted to the study of the nervous system and behavior. 2013;49(8):2007–20. doi:10.1016/j.cortex.2012.11.007

41. Mayes AR, Isaac CL, Holdstock JS, Cariga P, Gummer A, Roberts N. Long-term amnesia: a review and detailed illustrative case study. Cortex; a journal devoted to the study of the nervous system and behavior. 2003;39(4-5):567–603. doi:10.1016/s0010-9452(08)70855-4

42. Voltzenlogel V, Vignal J-P, Hirsch E, Manning L. The influence of seizure frequency on anterograde and remote memory in mesial temporal lobe epilepsy. Seizure. 2014;23(9):792–8. doi:10.1016/j.seizure.2014.06.013

43. Voltzenlogel V, Hirsch E, Vignal J-P, Valton L, Manning L. Preserved anterograde and remote memory in drug-responsive temporal lobe epileptic patients. Epilepsy research. 2015;115(ema, 8703089):126-32. doi:10.1016/j.eplepsyres.2015.06.006

44. Voltzenlogel V, Despres O, Vignal J-P, Steinhoff BJ, Kehrli P, Manning L. Remote memory in temporal lobe epilepsy. Epilepsia. 2006;47(8):1329–36. doi:10.1111/j.1528-1167.2006.00555.x

45. Borghesani V, Narvid J, Battistella G, et al. “Looks familiar, but I do not know who she is”: The role of the anterior right temporal lobe in famous face recognition. Cortex. 2019;115:72–85.

46. Drane DL, Ojemann JG, Phatak V, et al. Famous face identification in temporal lobe epilepsy: support for a multimodal integration model of semantic memory. Cortex; a journal devoted to the study of the nervous system and behavior. 2013;49(6):1648–67. doi:10.1016/j.cortex.2012.08.009

47. Seidenberg M, Griffith R, Sabsevitz D, et al. Recognition and identification of famous faces in patients with unilateral temporal lobe epilepsy. Neuropsychologia. 2002;40(4):446–56. doi:10.1016/s0028-3932(01)00096-3

48. Lah S, Grayson S, Lee T, Miller L. Memory for the past after temporal lobectomy: Impact of epilepsy and cognitive variables. Neuropsychologia. 2004;42(12):1666–1679. doi:10.1016/j.neuropsychologia.2004.04.008

49. Volfart A, Jonas J, Maillard L, Busigny T, Rossion B, Brissart H. Typical visual unfamiliar face individuation in left and right mesial temporal epilepsy. Neuropsychologia. 2020;147((Volfart, Jonas, Maillard, Rossion, Brissart) Universite de Lorraine, CNRS, CRAN, Nancy F-54000, France(Volfart, Busigny, Rossion) Universite Catholique de Louvain, Institute of Research in Psychological Science, Louvain-La-Neuve, Belgium(Jonas, Maillard):107583. doi:10.1016/j.neuropsychologia.2020.107583

50. Benke T, Kuen E, Schwarz M, Walser G. Proper name retrieval in temporal lobe epilepsy: naming of famous faces and landmarks. Epilepsy & behavior : E&B. 2013;27(2):371–7. doi:10.1016/j.yebeh.2013.02.013

51. Ashjazadeh N, Namjoo-Moghadam A, Mani A, et al. Comparison of executive function in idiopathic generalized epilepsy versus temporal lobe epilepsy. Neurocase. 2024;30(5):167–173.

52. Wang J, Xia X, Zhang B, et al. Association of glymphatic system dysfunction with cognitive impairment in Temporal lobe epilepsy. Frontiers in aging neuroscience. 2024;16:1459580.

53. Tedrus GMAS, Passos MLGA, Vargas LM, Menezes LEFJ. Cognition and epilepsy: Cognitive screening test. Dementia & Neuropsychologia. 2020;14(2):186–193. doi:10.1590/1980-57642020dn14-020013

54. Jaimes-Bautista AG, Rodriguez-Camacho M, Martinez-Juarez IE, Rodriguez-Agudelo Y. Quantitative and qualitative analysis of semantic verbal fluency in patients with temporal lobe epilepsy. Neurologia. 2020;35(1):1–9. doi:10.1016/j.nrl.2017.07.001

55. Kellermann TS, Bonilha L, Eskandari R, Garcia-Ramos C, Lin JJ, Hermann BP. Mapping the neuropsychological profile of temporal lobe epilepsy using cognitive network topology and graph theory. Epilepsy & Behavior. 2016;63(Alessio, A., Pereira, F.R., Sercheli, M.S., Rondina, J.M., Ozelo, H.B., Bilevicius, E., et al. (2013). Brain plasticity for verbal and visual memories in patients with mesial temporal lobe epilepsy and hippocampal sclerosis: an fMRI study. Hum Brain Mapp):9-16. doi:10.1016/j.yebeh.2016.07.030

56. Hwang G, Dabbs K, Conant L, et al. Cognitive slowing and its underlying neurobiology in temporal lobe epilepsy. Cortex. 2019;117((Hwang, Birn, Meyerand, Prabhakaran) Medical Physics, University of Wisconsin-Madison, Madison, WI, United States(Dabbs, Almane, Struck, Maganti, Hermann) Neurology, University of Wisconsin-Madison, Madison, WI, United States(Conant, Humphries, Raghavan):41-52. doi:10.1016/j.cortex.2019.02.022

57. Vanli Yavuz EN, Bilgic B, Matur Z, et al. Comparison of cognitive parameters between bilateral and unilateral hippocampal sclerosis. Noropsikiyatri Arsivi. 2016;53(3):199–204. doi:10.5152/npa.2016.14862

58. Struck AF, Garcia-Ramos C, Nair VA, et al. The relevance of Spearman’s g for epilepsy. Brain Communications. 2024;6(3):fcae176.

59. Jensen EJ, Hargreaves I, Bass A, Pexman P, Goodyear BG, Federico P. Cortical reorganization and reduced efficiency of visual word recognition in right temporal lobe epilepsy: a functional MRI study. Epilepsy research. 2011;93(2-3):155–63. doi:10.1016/j.eplepsyres.2010.12.003

60. Eichstaedt KE, Soble JR, Kamper JE, et al. Sex differences in lateralization of semantic verbal fluency in temporal lobe epilepsy. Brain and language. 2015;141(7506220, b5h):11-5. doi:10.1016/j.bandl.2014.11.013

61. Muller LC, Mader-Joaquim MJ, Terra VC, et al. Nonverbal fluency assessed by the five-point test in epilepsy patients with unilateral mesial temporal sclerosis-A Brazilian study. The Clinical Neuropsychologist. 2021;35(Suppl 1):S21–S31. doi:10.1080/13854046.2021.1887357

62. Poch C, Toledano R, Jimenez-Huete A, Garcia-Morales I, Gil-Nagel A, Campo P. Differences in visual naming performance between patients with temporal lobe epilepsy associated with temporopolar lesions versus hippocampal sclerosis. Neuropsychology. 2016;30(7):841–52. doi:10.1037/neu0000269

63. Tudesco IdSS, Vaz LJ, Mantoan MAS, et al. Assessment of working memory in patients with mesial temporal lobe epilepsy associated with unilateral hippocampal sclerosis. Epilepsy & behavior : E&B. 2010;18(3):223–8. doi:10.1016/j.yebeh.2010.04.021

64. Nagele M, Yetkin Z, Bailey KC, et al. Non-language neuropsychological measures increase sensitivity of identifying language reorganization in patients with epilepsy: a pilot study. Journal of Clinical and Experimental Neuropsychology. 2024;46(10):978–988.

65. Lah S, Lee T, Grayson S, Miller L. Changes in retrograde memory following temporal lobectomy. Epilepsy and Behavior. 2008;13(2):391–396. doi:10.1016/j.yebeh.2008.05.002

66. Saykin AJ, Stafiniak P, Robinson LJ, et al. Language before and after temporal lobectomy: Specificity of acute changes and relation to early risk factors. Epilepsia. 1995;36(11):1071–1077. doi:10.1111/j.1528-1157.1995.tb00464.x

67. Sroubek J, Kramska L, Nova M, et al. Multiple Hippocampal Transections: Initial Clinical Experience with Modified Technique. World Neurosurgery. 2025;196((Sroubek, Klener) Department of Neurosurgery, Na Homolce Hospital, Prague, Czechia(Sroubek, Cesak) Department of Neurosurgery, Faculty of Medicine, Charles University, Hradec Kralove, Czechia(Kramska) Department of Clinical Psychology, Na Homolce Hospital):123804. doi:10.1016/j.wneu.2025.123804

68. Martin RC, Loring DW, Meador KJ, Lee GP. The effects of lateralized temporal lobe dysfunction on formal and semantic word fluency. Neuropsychologia. 1990;28(8):823–9. doi:10.1016/0028-3932(90)90006-a

69. Perrone-Bertolotti M, Zoubrinetzky R, Yvert G, Le Bas JF, Baciu M. Functional MRI and neuropsychological evidence for language plasticity before and after surgery in one patient with left temporal lobe epilepsy. Epilepsy & behavior : E&B. 2012;23(1):81–6. doi:10.1016/j.yebeh.2011.11.011

70. Prayson BE, Prayson RA, Kubu CS, Bingaman W, Najm IM, Busch RM. Effects of dual pathology on cognitive outcome following left anterior temporal lobectomy for treatment of epilepsy. Epilepsy & Behavior. 2013;28(3):426–431. doi:10.1016/j.yebeh.2013.05.040

71. de Souza JPSAS, Ayub G, Nogueira M, et al. Temporopolar amygdalohippocampectomy: Seizure control and postoperative outcomes. Journal of Neurosurgery. 2021;134(4):1044–1053. doi:10.3171/2020.3.JNS192624

72. Du Preez K. Symptomatic and functional concomitants of anterior temporal lobe surgery. Dissertation Abstracts International: Section B: The Sciences and Engineering. 2022;83(3-B):No-Specified.

73. Šroubek J, Krámská L, Nová M, et al. Multiple Hippocampal Transections: Initial Clinical Experience with Modified Technique. World Neurosurgery. 2025;196:123804.

74. Bartha L, Trinka E, Ortler M, et al. Linguistic deficits following left selective amygdalohippocampectomy: a prospective study. Epilepsy & behavior : E&B. 2004;5(3):348–57. doi:10.1016/j.yebeh.2004.02.004

75. Giovagnoli AR, Parente A, Didato G, et al. The course of language functions after temporal lobe epilepsy surgery: a prospective study. European journal of neurology. 2016;23(12):1713–1721. doi:10.1111/ene.13113

76. Yang P-F, Zhang H-J, Pei J-S, et al. Neuropsychological outcomes of subtemporal selective amygdalohippocampectomy via a small craniotomy. Journal of neurosurgery.2016;125(1):67–74. doi:10.3171/2015.6.JNS1583

77. Howard D, Patterson KE. The pyramids and palm trees test. 1992;

78. Bozeat S, Ralph MAL, Patterson K, Garrard P, Hodges JR. Non-verbal semantic impairment in semantic dementia. Neuropsychologia. 2000;38(9):1207–1215.

79. Manes F, Hodges JR, Graham KS, Zeman A. Focal autobiographical amnesia in association with transient epileptic amnesia. Brain : a journal of neurology. 2001;124(Pt 3):499–509. doi:10.1093/brain/124.3.499

80. Giovagnoli AR, Bell B. Drawing from memory in focal epilepsy: a cognitive and neural perspective. Epilepsy research. 2011;94(1-2):69–74. doi:10.1016/j.eplepsyres.2011.01.004

81. Aleman-Gomez Y, Poch C, Toledano R, et al. Morphometric correlates of anomia in patients with small left temporopolar lesions. Journal of neuropsychology. 2020;14(2):260–282. doi:10.1111/jnp.12184

82. Campo P, Poch C, Toledano R, et al. Visual object naming in patients with small lesions centered at the left temporopolar region. Brain structure & function. 2016;221(1):473–85. doi:10.1007/s00429-014-0919-1

83. Toledano R, Jimenez-Huete A, Garcia-Morales I, et al. Aphasic seizures in patients with temporopolar and anterior temporobasal lesions: a video-EEG study. Epilepsy & behavior : E&B. 2013;29(1):172–7. doi:10.1016/j.yebeh.2013.07.017

84. Poch C, Toledano R, Garcia-Morales I, Aleman-Gomez Y, Gil-Nagel A, Campo P. Contributions of left and right anterior temporal lobes to semantic cognition: Evidence from patients with small temporopolar lesions. Neuropsychologia. 2021;152((Poch) Facultad de Lenguas y Educacion, Universidad Nebrija, Spain(Toledano, Garcia-Morales, Gil-Nagel) Hospital Ruber Internacional, Epilepsy Unit, Neurology Department, Madrid, Spain(Toledano) University Hospital of Ramon y Cajal, Epilepsy Unit, Neurolo):107738. doi:10.1016/j.neuropsychologia.2020.107738

85. Rice GE, Caswell H, Moore P, Lambon Ralph MA, Hoffman P. Revealing the Dynamic Modulations That Underpin a Resilient Neural Network for Semantic Cognition: An fMRI Investigation in Patients With Anterior Temporal Lobe Resection. Cerebral cortex (New York, NY : 1991). 2018;28(8):3004-3016. doi:10.1093/cercor/bhy116

86. Collins AM, Loftus EF. A spreading-activation theory of semantic processing. Psychological review. 1975;82(6):407.

87. Blaxton TA, Bookheimer SY. Retrieval inhibition in anomia. Brain and language. 1993;44(2):221–37. doi:10.1006/brln.1993.1015

88. Miyamoto T, Kohsaka M, Katayama J, et al. Evaluation of semantic processing in patients with temporal lobe epilepsy using event-related potential. Psychiatry and clinical neurosciences. 1995;49(3):S237–9. doi:10.1111/j.1440-1819.1995.tb02188.x

89. Miozzo M, Hamberger MJ. Preserved meaning in the context of impaired naming in temporal lobe epilepsy. Neuropsychology. 2015;29(2):274–281. doi:10.1037/neu0000097

90. Lechowicz M, Miller L, Irish M, Addis DR, Mohamed A, Lah S. Imagining future events in patients with unilateral temporal lobe epilepsy. The British journal of clinical psychology. 2016;55(2):187–205. doi:10.1111/bjc.12107

91. St-Laurent M, Moscovitch M, Levine B, McAndrews MP. Determinants of autobiographical memory in patients with unilateral temporal lobe epilepsy or excisions. Neuropsychologia. 2009;47(11):2211–21. doi:10.1016/j.neuropsychologia.2009.01.032

92. Viskontas IV, McAndrews MP, Moscovitch M. Remote episodic memory deficits in patients with unilateral temporal lobe epilepsy and excisions. The Journal of neuroscience :the official journal of the Society for Neuroscience. 2000;20(15):5853–7.

93. Herfurth K, Kasper B, Schwarz M, Stefan H, Pauli E. Autobiographical memory in temporal lobe epilepsy: role of hippocampal and temporal lateral structures. Epilepsy & behavior : E&B. 2010;19(3):365–71. doi:10.1016/j.yebeh.2010.07.012

94. Rayner G, Siveges B, Allebone J, Pieters J, Wilson SJ. Contribution of autobiographic memory impairment to subjective memory complaints in focal epilepsy. Epilepsy & behavior : E&B. 2020;102(100892858):106636. doi:10.1016/j.yebeh.2019.106636

95. Munera CP, Lomlomdjian C, Gori B, et al. Episodic and semantic autobiographical memory in temporal lobe epilepsy. Epilepsy Research and Treatment. 2014;2014((Munera, Lomlomdjian, Gori, Terpiluk, Medel, Solis, Kochen) Epilepsy Center, Neurology Division, Ramos Mejia Hospital, Gral Urquiza 609, Buenos Aires C1221ADC, Argentina(Munera, Lomlomdjian, Gori, Terpiluk, Medel, Solis, Kochen) Center for Clinical and Ex):157452. doi:10.1155/2014/157452

96. Blaxton TA. Dissociations among memory measures in memory-impaired subjects: evidence for a processing account of memory. Memory & cognition. 1992;20(5):549–62. doi:10.3758/bf03199587

97. Delazer M, Gasperi A, Bartha L, Trinka E, Benke T. Number processing in temporal lobe epilepsy. Journal of neurology, neurosurgery, and psychiatry. 2004;75(6):901–3. doi:10.1136/jnnp.2003.023614

98. Messas CS, Mansur LL, Castro LHM. Semantic memory impairment in temporal lobe epilepsy associated with hippocampal sclerosis. Epilepsy & behavior : E&B. 2008;12(2):311–6. doi:10.1016/j.yebeh.2007.10.014

99. Miro J, Gurtubay-Antolin A, Ripolles P, et al. Interhemispheric microstructural connectivity in bitemporal lobe epilepsy with hippocampal sclerosis. Cortex: A Journal Devoted to the Study of the Nervous System and Behavior. 2015;67(Adam, C. (2006). How do the temporal lobes communicate in medial temporal lobe seizures? Revue Neurologique, 162(8-9), 813-818. https://pubmed.ncbi.nlm.nih.gov/17028541https://dx.doi.org/10.1016/S0035-3787(06)75083-4 2006-21639-002.Adam, C., Hasboun, D.):106-121. doi:10.1016/j.cortex.2015.03.018

100. Poprelka K, Patrikelis P, Takousi M, et al. Arousal deregulation in the co-shaping of neuropsychological dysfunction in frontal and mesial temporal lobe epilepsy. Epilepsy Research. 2023;194((Poprelka, Patrikelis, Fasilis, Margariti, Ntinopoulou, Verentzioti, Stefanatou, Alexoudi, Korfias, Gatzonis) 1st Department of Neurosurgery, National & Kapodistrian University of Athens, Greece(Patrikelis, Messinis) Laboratory of Cognitive Neuroscience):107189. doi:10.1016/j.eplepsyres.2023.107189

101. Abony-Bencze K. Memory deficits before and after unilateral anterior temporal lobectomy: A comparative study of children and adults. Dissertation Abstracts International: Section B: The Sciences and Engineering. 1999;60(6-B):2970.

102. Zannino GD, Murolo R, Grammaldo L, et al. Visuo-verbal distinction revisited: new insights from a study on temporal lobe epilepsy patients in the debate over the lateralization of material-specific and process-specific aspects of memory. Journal of clinical and experimental neuropsychology. 2020;42(10):1085–1098. doi:10.1080/13803395.2020.1844868

103. Metternich B, Buschmann F, Wagner K, Schulze-Bonhage A, Kriston L. Verbal fluency in focal epilepsy: a systematic review and meta-analysis. Neuropsychology review. 2014;24:200–218.

104. Henry JD, Crawford JR. A meta-analytic review of verbal fluency performance in patients with traumatic brain injury. Neuropsychology. 2004;18(4):621.

105. Gleissner U, Elger CE. The hippocampal contribution to verbal fluency in patients with temporal lobe epilepsy. Cortex; a journal devoted to the study of the nervous system and behavior. 2001;37(1):55–63. doi:10.1016/s0010-9452(08)70557-4

106. Baldo JV, Schwartz S, Wilkins D, Dronkers NF. Role of frontal versus temporal cortex in verbal fluency as revealed by voxel-based lesion symptom mapping. Journal of the International Neuropsychological Society. 2006;12(6):896–900.

107. Rossion B, Caldara R, Seghier M, Schuller AM, Lazeyras F, Mayer E. A network of occipitolJtemporal facelJsensitive areas besides the right middle fusiform gyrus is necessary for normal face processing. Brain. 2003;126(11):2381–2395.

108. Sodov U, Avirmed T, Zuunnast K. Cognitive Impairment of Temporal Lobe Epilepsy and Risk Factors. Journal of Neurology & Neuropsychiatry. 2024;1:17–25.

109. Collins JA, Olson IR. Beyond the FFA: the role of the ventral anterior temporal lobes in face processing. Neuropsychologia. 2014;61:65–79.

110. Helmstaedter C, Elger C, Vogt V. Cognitive outcomes more than 5 years after temporal lobe epilepsy surgery: Remarkable functional recovery when seizures are controlled. Seizure. 2018;62:116–123.

111. Drane DL, Loring DW, Voets NL, et al. Better object recognition and naming outcome with MRI-guided stereotactic laser amygdalohippocampotomy for temporal lobe epilepsy. Epilepsia. 2015;56(1):101–13. doi:10.1111/epi.12860

112. Lezak MD. Neuropsychological assessment. Oxford University Press, USA; 2004.

113. Behrmann M, Plaut DC. Bilateral hemispheric processing of words and faces: evidence from word impairments in prosopagnosia and face impairments in pure alexia. Cerebral Cortex. 2014;24(4):1102–1118.

114. Pobric G, Ralph MAL, Jefferies E. The role of the anterior temporal lobes in the comprehension of concrete and abstract words: rTMS evidence. Cortex. 2009;45(9):1104–1110.

115. Pobric G, Jefferies E, Ralph MAL. Anterior temporal lobes mediate semantic representation: mimicking semantic dementia by using rTMS in normal participants. Proceedings of the National Academy of Sciences. 2007;104(50):20137–20141.

116. Helmstaedter C. Cognitive outcomes of different surgical approaches in temporal lobe epilepsy. Epileptic disorders. 2013;15:221–239.

