## Supplementary material for "Semantic Functioning in Temporal Lobe Epilepsy: A Systematic Review and Meta-Analysis": Table 1

Table 1. Summary results of main and sub-group analyses.

| Comparison | | | *k* | *m* | *g* ^a^ | 95% CI | | Total I² (%) |
| --- | --- | --- | --- | --- | --- | --- | --- | --- |
| **Semantic Fluency** | | | | | | |  |  |
| *Any Semantic Category* | | | |  |  |  | |  |
|  | HC vs TLE | | 42 | 117 | -1.35 | **-1.67, -1.04** | | 99.86 |
|  |  | HC vs mixed TLE | 20 | 34 | -1.70 | **-2.34, -1.06** | | 99.72 |
|  |  | HC vs RTLE | 18 | 35 | -1.00 | **-1.21, -0.80** | | 97.36 |
|  |  | HC vs LTLE | 23 | 49 | -1.17 | **-1.44, -0.90** | | 99.11 |
|  | RTLE vs LTLE | | 28 | 51 | -0.25 | **-0.39, -0.11** | | 97.38 |
|  | Pre vs Post Surgery | | 8 | 25 | -0.07 | -0.36, 0.23 | | 98.93 |
|  |  | Mixed TLE | 2 | 3 | -0.07 | -0.66, 0.51 | | 95.60 |
|  |  | RTLE | 6 | 11 | -0.15 | -0.72, 0.41 | | 99.03 |
|  |  | LTLE | 6 | 11 | -0.03 | -0.33, 0.28 | | 93.15 |
| *Animals* | | |  |  |  |  | |  |
|  | HC vs TLE | | 22 | 27 | -1.17 | **-1.46, -0.88** | | 98.68 |
|  |  | HC vs mixed TLE | 8 | 8 | -1.02 | **-1.47, -0.56** | | 96.98 |
|  |  | HC vs RTLE | 7 | 7 | -0.93 | **-1.23, -0.63** | | 61.80 |
|  |  | HC vs LTLE | 12 | 12 | -1.30 | **-1.82, -0.79** | | 98.08 |
|  | RTLE vs LTLE | | 13 | 18 | -0.33 | **-0.53, -0.12** | | 91.51 |
|  | Pre vs Post Surgery | | 5 | 11 | -0.05 | -0.54, 0.43 | | 98.69 |
|  |  | Mixed TLE | 2 | 3 | -0.07 | -0.66, 0.51 | | 95.60 |
|  |  | RTLE | 3 | 4 | -0.16 | -1.17, 0.86 | | 98.24 |
|  |  | LTLE | 3 | 4 | -0.05 | -0.70, 0.61 | | 94.22 |
| **WAIS-IV Vocabulary** | | | | | | |  |  |
|  | HC vs TLE | | 3 | 2 | -1.08 | **-1.57, -0.58** | | 33.32 |
|  |  | HC vs mixed TLE | 1 | 1 | -1.12 | -2.10, -0.19 | | n.a. |
|  |  | HC vs RTLE | 1 | 1 | -1.44 | **-2.21, -0.69** | | n.a. |
|  |  | HC vs LTLE | 1 | 1 | -0.69 | -1.41, 0.02 | | n.a. |
|  | RTLE vs LTLE | | 3 | 4 | -0.16 | -0.48, 0.17 | | 3.63 |
| **PPT** | | | | | | |  |  |
|  | HC vs TLE | | 3 | 4 | -0.44 | -1.50, 0.61 | | 86.34 |
|  |  | HC vs mixed TLE | n.a. | n.a. | n.a. | n.a. | | n.a. |
|  |  | HC vs RTLE | n.a. | n.a. | n.a. | n.a. | | n.a. |
|  |  | HC vs LTLE | 1 | 1 | -1.41 | **-2.35, -0.47** | | n.a. |
|  | RTLE vs LTLE | | n.a. | n.a. | n.a. | n.a. | | n.a. |
| **CCT** | | | | | | |  |  |
|  | HC vs TLE | | 5 | 7 | -1.37 | **-1.76, -0.99** | | 74.69 |
|  |  | HC vs mixed TLE | 3 | 4 | -1.17 | **-1.53, -0.81** | | 0.00 |
|  |  | HC vs RTLE | 1 | 1 | -1.96 | **-2.74,  -1.17** | | n.a. |
|  |  | HC vs LTLE | 2 | 2 | -1.37 | **-1.75, -0.99** | | 56.11 |
|  | RTLE vs LTLE | | n.a. | n.a. | n.a. | n.a. | | n.a. |

^a^ Negative values indicate poorer performance for the second named condition.

*k* = number of studies, m = number of effect sizes, g = standardised mean difference (Hedges’ g), CI = confidence interval, I2 = proportion of systematic statistical heterogeneity, HC = healthy controls, TLE = temporal lobe epilepsy, LTLE = left temporal lobe epilepsy, RTLE = right temporal lobe epilepsy, WAIS-IV = Weschler Abbreviated Intelligence Scale – 4^th^ Edition, PPT = Pyramids and Palm Trees, CCT = Camels and Cactus Test.

Confidence intervals that do not cross zero are presented in bold to indicate statistical significance.
