## Supplementary material for "Semantic Functioning in Temporal Lobe Epilepsy: A Systematic Review and Meta-Analysis": Figure 1

Figure 1. PRISMA Workflow of Literature Screening

**Identification**

Studies screened **(*n* = 1153)**

Studies assessed for eligibility **(*n* = 389)**

References removed **(*n* = 888)**

Duplicates identified manually (*n* = 12)

Duplicates identified by Covidence (*n* = 876)

Studies excluded **(*n* = 764)**

Studies excluded **(*n* = 248)**

Not in English (*n* = 4)

Incorrect outcomes (*n* = 87)

Paediatric population (*n* = 1)

Wrong patient population (*n* = 72)

Unclear epilepsy diagnosis (*n* = 28)

Not original data (*n* = 4)

Full text not available (*n* = 52)

**Included**

Studies included in review **(*N* = 141)**

**Screening**

Studies from databases/registers **(*n* = 2041)**

Embase (*n* = 853)

PsycINFO (*n* = 625)

MEDLINE (*n* = 544)

Google Scholar (*n* = 19)

*Note*. PRISMA = Preferred Reporting for Systematic Reviews and Meta-Analysis.
