## Supplementary material for "Semantic Functioning in Temporal Lobe Epilepsy: A Systematic Review and Meta-Analysis": Figure 2

Figure 2. Study Quality Analysis Using the Newcastle-Ottawa Quality Assessment Scale

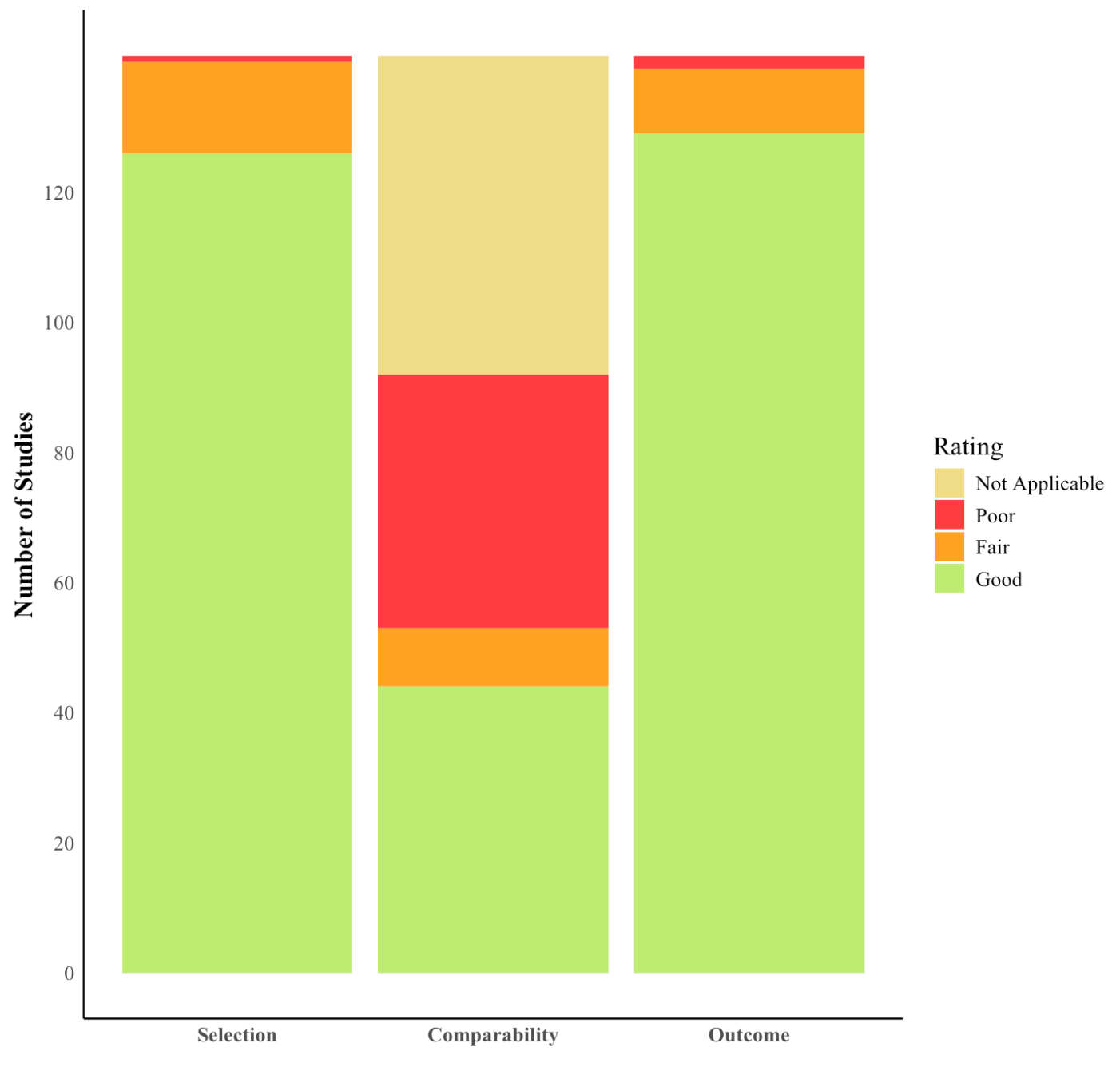

*Note*. The column colours represent the study's overall risk of bias across the domains of Selection, Comparability, and Outcomes: (i) red indicates high risk of bias, (ii) yellow indicates medium or unclear risk of bias, and (iii) green indicates low risk of bias. The adjusted thresholds for converting the Newcastle-Ottawa scales to AHRQ standards are based on the percentage of stars achieved: Selection is rated as good (≥ 75%), fair (50-74%), or poor (< 50%); Comparability is rated as good (100%), fair (50%) or poor (0%); Outcome is rated as good (≥ 67%), fair (33-66%), or poor (< 33%).
