## Supplementary material for "Semantic Functioning in Temporal Lobe Epilepsy: A Systematic Review and Meta-Analysis": Figure 3

Figure 3. Lateralisation of Semantic Memory Function in Temporal Lobe Epilepsy (TLE): Summary of Empirical Evidence

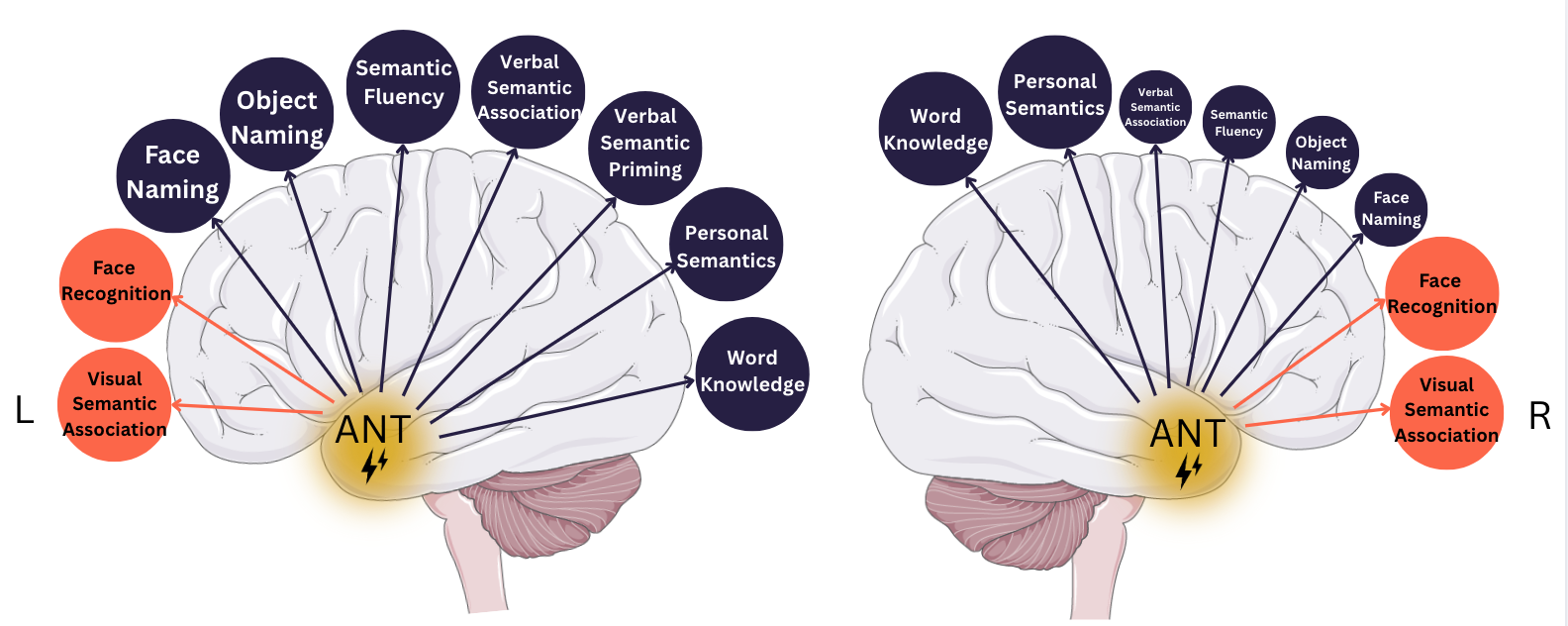

*Note*. AmNT = anterior mesial temporal lobe. Circle colour represents input modality: navy = verbal measures, orange = visual measures. Smaller circles indicate functions that are less impaired relative to the contralateral hemisphere.
