## Supplementary Table 2 for "Semantic Functioning in Temporal Lobe Epilepsy: A Systematic Review and Meta-Analysis"

| Supplementary Table 2. Quality ratings on the modified NOS for included studies | | | | |  |  | |  |  | | | |  |
| --- | --- | --- | --- | --- | --- | --- | --- | --- | --- | --- | --- | --- | --- |
|  | Selection | | | |  | Comparability | |  | Outcome | | | |  |
| Study | Representativeness of the TLE cohort | Selection of non-  exposed cohort^a^ | Ascertainment of TLE diagnosis | Total |  | Comparability of cohorts^a^ | Total |  | Formal assessment of semantic memory | Sufficient follow-up length^a^ | Adequacy of cohort follow-up^a^ | Total | # (%) of stars out of total eligible |
| Abony-Bencze, 1999 | * | n/a | - | 1/2 |  | -- | 0/2 |  | * | * | * | 3/3 | 4/5 (80) |
| Alemán‐Gómez et al., 2020 | * | * | * | 3/3 |  | ** | 2/2 |  | * | - | n/a | 1/2 | 6/7 (86) |
| Antonucci et al., 2008 | * | * | * | 3/3 |  | ** | 2/2 |  | * | * | n/a | 2/2 | 7/7 (100) |
| Ashjazadeh et al., 2024 | * | * | * | 3/3 |  | *- | 1/2 |  | * | n/a | n/a | 1/1 | 5/6 (83) |
| Avila et al., 2001 | * | n/a | * | 2/2 |  | n/a | n/a |  | * | n/a | n/a | 1/1 | 3/3 (100) |
| Barnett et al., 2014 | * | n/a | * | 2/2 |  | n/a | n/a |  | * | n/a | n/a | 1/1 | 3/3 (100) |
| Barrett Jones et al., 2022 | * | n/a | * | 2/2 |  | n/a | n/a |  | * | n/a | n/a | 1/1 | 3/3 (100) |
| Bartha et al., 2004 | * | n/a | * | 2/2 |  | n/a | n/a |  | * | * | * | 3/3 | 5/5 (100) |
| Bell et al., 2001 | * | * | * | 3/3 |  | -- | 0/2 |  | * | n/a | n/a | 1/1 | 4/6 (67) |
| Benke et al., 2013 | * | * | * | 3/3 |  | ** | 2/2 |  | * | n/a | n/a | 1/1 | 6/6 (100) |
| Bidet-Caulet et al., 2009 | * | * | * | 3/3 |  | ** | 2/2 |  | * | * | - | 2/3 | 7/8 (88) |
| Billingsley et al., 2000 | * | - | * | 2/3 |  | -- | 0/2 |  | * | n/a | n/a | 1/1 | 3/6 (50) |
| Blackmon et al., 2019 | * | * | * | 3/3 |  | -- | 0/2 |  | * | n/a | n/a | 1/1 | 4/6 (67) |
| Blaxton et al., 1993 | * | * | * | 3/3 |  | -- | 0/2 |  | * | - | n/a | 1/2 | 4/7 (57) |
| Blaxton, 1992 | * | * | * | 3/3 |  | -- | 0/2 |  | * | - | n/a | 1/2 | 4/7 (57) |
| Bochyńska et al., 2023 | * | * | * | 3/3 |  | ** | 2/2 |  | * | n/a | n/a | 1/1 | 6/6 (100) |
| Caciagli et al., 2023 | * | * | * | 3/3 |  | *- | 1/2 |  | * | n/a | n/a | 1/1 | 5/6 (83) |
| Cairós-González et al., 2024 | * | * | * | 3/3 |  | -- | 0/2 |  | * | n/a | n/a | 1/1 | 4/6 (67) |
| Campo et al., 2016 | * | * | * | 3/3 |  | ** | 2/2 |  | * | n/a | n/a | 1/1 | 6/6 (100) |
| Chang et al., 2017 | * | * | * | 3/3 |  | -- | 0/2 |  | * | n/a | n/a | 1/1 | 4/6 (67) |
| Condret-Santi et al., 2014 | * | * | * | 3/3 |  | ** | 2/2 |  | * | - | n/a | 1/2 | 6/7 (86) |
| de Souza et al., 2021 | * | n/a | * | 2/2 |  | n/a | n/a |  | * | * | - | 2/3 | 4/5 (80) |
| Delazer et al., 2004 | * | * | * | 3/3 |  | ** | 2/2 |  | * | n/a | n/a | 1/1 | 6/6 (100) |
| Doherty et al., 2021 | * | n/a | * | 2/2 |  | n/a | n/a |  | * | n/a | n/a | 1/1 | 3/3 (100) |
| Drane et al., 2006 | * | n/a | * | 2/2 |  | n/a | n/a |  | * | n/a | n/a | 1/1 | 3/3 (100) |
| Drane et al., 2008 | * | n/a | * | 2/2 |  | n/a | n/a |  | * | * | n/a | 2/2 | 4/4 (100) |
| Drane et al., 2013 | * | * | * | 3/3 |  | -- | 0/2 |  | * | * | * | 3/3 | 6/8 (75) |
| Drane et al., 2015 | * | n/a | * | 2/2 |  | n/a | n/a |  | * | * | * | 3/3 | 5/5 (100) |
| Drane et al., 2016 | * | n/a | * | 2/2 |  | n/a | n/a |  | * | n/a | n/a | 1/1 | 3/3 (100) |
| Dredla et al., 2016 | * | n/a | * | 2/2 |  | n/a | n/a |  | * | * | n/a | 2/2 | 4/4 (100) |
| Du Preez, 2020 | * | n/a | * | 2/2 |  | n/a | n/a |  | * | * | * | 3/3 | 5/5 (100) |
| Eichstaedt et al., 2015 | * | n/a | * | 2/2 |  | n/a | n/a |  | * | n/a | n/a | 1/1 | 3/3 (100) |
| Ellis et al.,1989 | * | - | * | 2/3 |  | -- | 0/2 |  | * | * | * | 3/3 | 5/8 (63) |
| Ferrario et al., 2024 | * | * | * | 3/3 |  | -- | 0/2 |  | * | n/a | n/a | 1/1 | 4/6 (67) |
| Flugel et al., 2006 | * | n/a | * | 2/2 |  | n/a | n/a |  | * | n/a | n/a | 1/1 | 3/3 (100) |
| Foesleitner et al., 2021 | * | n/a | * | 2/2 |  | n/a | n/a |  | * | * | - | 2/3 | 4/5 (80) |
| Forthoffer et al., 2025 | * | n/a | * | 2/2 |  | n/a | n/a |  | * | - | * | 2/3 | 4/5 |
| Frank et al., 2018 | * | n/a | * | 2/2 |  | n/a | n/a |  | * | n/a | n/a | 1/1 | 3/3 (100) |
| Galioto et al., 2023 | * | * | * | 3/3 |  | -- | 0/2 |  | * | n/a | n/a | 1/1 | 4/6 (67) |
| Giovagnoli & Bell, 2011 | * | * | * | 3/3 |  | ** | 2/2 |  | * | n/a | n/a | 1/1 | 6/6 (100) |
| Giovagnoli et al., 1999 | * | * | * | 3/3 |  | -- | 0/2 |  | * | n/a | n/a | 1/1 | 4/6 (67) |
| Giovagnoli, 2005 | * | * | * | 3/3 |  | -- | 0/2 |  | * | n/a | n/a | 1/1 | 4/6 (67) |
| Giovagnoli et al., 2005 | * | * | * | 3/3 |  | -- | 0/2 |  | * | n/a | n/a | 1/1 | 4/6 (67) |
| Giovagnoli et al., 2009 | * | - | * | 2/3 |  | -- | 0/2 |  | * | n/a | n/a | 1/1 | 3/6 (50) |
| Giovagnoli et al., 2016 | * | * | * | 3/3 |  | -- | 0/2 |  | * | * | * | 3/3 | 6/8 (75) |
| Giovagnoli, 2020 | * | * | * | 3/3 |  | -- | 0/2 |  | * | n/a | n/a | 1/1 | 4/6 (67) |
| Giovannetti et al., 2003 | * | * | * | 3/3 |  | ** | 2/2 |  | * | n/a | n/a | 1/1 | 6/6 (100) |
| Gleissner & Elger, 2001 | * | * | * | 3/3 |  | *- | 1/2 |  | * | n/a | n/a | 1/1 | 5/6 (83) |
| Griffith et al., 2006 | * | * | * | 3/3 |  | -- | 0/2 |  | * | n/a | n/a | 1/1 | 4/6 (67) |
| Herfurth et al., 2010 | * | * | * | 3/3 |  | ** | 2/2 |  | * | - | n/a | 1/2 | 6/7 (86) |
| Hermann et al., 1995 | * | n/a | * | 2/2 |  | n/a | n/a |  | * | * | * | 3/3 | 5/5 (100) |
| Hwang et al., 2019 | * | * | * | 3/3 |  | -- | 0/2 |  | * | n/a | n/a | 1/1 | 4/6 (67) |
| Jaimes-Bautista et al., 2020 | * | * | * | 3/3 |  | ** | 2/2 |  | * | n/a | n/a | 1/1 | 6/6 (100) |
| Jensen et al., 2011a | * | * | * | 3/3 |  | -- | 0/2 |  | * | n/a | n/a | 1/1 | 4/6 (67) |
| Jensen et al., 2011b | * | * | * | 3/3 |  | ** | 2/2 |  | * | n/a | n/a | 1/1 | 6/6 (100) |
| Jokeit et al., 1998 | * | n/a | * | 2/2 |  | n/a | n/a |  | * | * | n/a | 2/2 | 4/4 (100) |
| Kellermann 2016 | * | * | * | 3/3 |  | -- | 0/2 |  | * | n/a | n/a | 1/1 | 4/6 (67) |
| Lah et al., 2004 | * | * | * | 3/3 |  | -- | 0/2 |  | * | * | * | 3/3 | 5/7 (71) |
| Lah et al., 2008 | * | n/a | * | 2/3 |  | n/a | n/a |  | * | * | * | 3/3 | 5/5 (100) |
| Lambon Ralph et al., 2012 | * | - | * | 2/3 |  | -- | 0/2 |  | * | * | n/a | 2/2 | 4/7 (57) |
| Lechowicz et al., 2016 | * | * | * | 3/3 |  | -- | 0/2 |  | * | - | n/a | 1/2 | 4/7 (57) |
| Lehéricy et al., 2000 | * | * | * | 3/3 |  | ** | 2/2 |  | * | n/a | n/a | 1/1 | 6/6 (100) |
| Lomlomdjian et al., 2011 | * | n/a | * | 2/2 |  | n/a | n/a |  | * | n/a | n/a | 1/1 | 3/3 (100) |
| Lomlomdjian et al., 2017 | * | n/a | * | 2/2 |  | n/a | n/a |  | * | n/a | n/a | 1/1 | 3/3 (100) |
| Longo et al., 2021 | * | n/a | * | 2/2 |  | n/a | n/a |  | * | - | n/a | 1/2 | 3/4 (75) |
| Luckhurst & Lloyd-Jones, 2001 | * | - | * | 2/3 |  | -- | 0/2 |  | * | * | n/a | 2/2 | 4/7 (57) |
| Manes et al., 2001 | - | * | * | 2/3 |  | ** | 2/2 |  | * | n/a | n/a | 1/1 | 5/6 (83) |
| Manning et al., 2013 | * | * | * | 3/3 |  | ** | 2/2 |  | * | * | n/a | 2/2 | 7/7 (100) |
| Martin et al., 1990 | * | * | * | 3/3 |  | -- | 0/2 |  | * | - | * | 2/3 | 5/8 (63) |
| Mayes et al., 2003 | * | * | * | 3/3 |  | ** | 2/2 |  | * | n/a | n/a | 1/1 | 6/6 (100) |
| Messas et al., 2008 | * | * | * | 3/3 |  | ** | 2/2 |  | * | n/a | n/a | 1/1 | 6/6 (100) |
| Miatton et al., 2011 | * | n/a | * | 2/2 |  | n/a | n/a |  | * | - | n/a | 1/2 | 3/4 (75) |
| Milton et al., 2010 | * | * | * | 3/3 |  | ** | 2/2 |  | * | n/a | n/a | 1/1 | 6/6 (100) |
| Miozzo & Hamberger, 2015 | * | * | * | 3/3 |  | ** | 2/2 |  | * | n/a | n/a | 1/1 | 6/6 (100) |
| Miró et al., 2015 | * | * | * | 3/3 |  | ** | 2/2 |  | * | n/a | n/a | 1/1 | 6/6 (100) |
| Miyamoto et al., 1995 | * | * | - | 2/3 |  | *- | 1/2 |  | * | n/a | n/a | 1/1 | 4/6 (67) |
| Miyamoto et al., 2000 | - | - | * | 1/3 |  | -- | 0/2 |  | * | - | - | 1/3 | 2/6 (33) |
| Muller et al., 2021 | * | n/a | * | 2/2 |  | n/a | n/a |  | * | n/a | n/a | 1/1 | 3/3 (100) |
| Múnera et al., 2014 | * | * | * | 3/3 |  | ** | 2/2 |  | * | n/a | n/a | 1/1 | 6/6 (100) |
| Nagele et al., 2024 | * | n/a | * | 2/2 |  | n/a | n/a |  | * | n/a | n/a | 1/1 | 3/3 (100) |
| N'Kaoua et al., 2001 | * | * | * | 3/3 |  | ** | 2/2 |  | * | n/a | n/a | 1/1 | 6/6 (100) |
| Osorio et al., 2017 | * | * | * | 3/3 |  | ** | 2/2 |  | * | n/a | n/a | 1/1 | 6/6 (100) |
| Parente et al., 2013 | * | * | * | 3/3 |  | -- | 0/2 |  | * | n/a | n/a | 1/1 | 4/6 (67) |
| Perrone-Bertolotti et al., 2012 | * | n/a | * | 2/2 |  | n/a | n/a |  | * | * | n/a | 2/2 | 4/4 (100) |
| Poch et al., 2016 | * | * | * | 3/3 |  | ** | 2/2 |  | * | n/a | n/a | 1/1 | 6/6 (100) |
| Poch et al., 2021 | * | * | * | 3/3 |  | -- | 0/2 |  | * | n/a | n/a | 1/1 | 4/6 (67) |
| Pope et al., 2019 | * | n/a | * | 2/2 |  | n/a | n/a |  | * | - | - | 1/1 | 3/3 (100) |
| Poprelka et al., 2023 | * | * | * | 3/3 |  | ** | 2/2 |  | * | n/a | n/a | 1/1 | 6/6 (100) |
| Prayson et al., 2013 | * | n/a | * | 2/2 |  | n/a | n/a |  | * | * | n/a | 2/2 | 4/4 (100) |
| Rayner et al., 2020 | * | * | * | 3/3 |  | -- | 0/2 |  | * | n/a | n/a | 1/1 | 4/6 (67) |
| Realmuto et al., 2015 | * | * | * | 3/3 |  | ** | 2/2 |  | * | n/a | n/a | 1/1 | 6/6 (100) |
| Reyes et al., 2021 | - | * | * | 2/3 |  | ** | 2/2 |  | * | n/a | n/a | 1/1 | 5/6 (83) |
| Rice et al., 2018a | * | * | * | 3/3 |  | *- | 1/2 |  | * | * | n/a | 2/2 | 6/7 (86) |
| Rice et al., 2018b | * | * | * | 3/3 |  | *- | 1/2 |  | * | * | n/a | 2/2 | 6/7 (86) |
| Roger et al., 2019 | * | n/a | * | 2/2 |  | n/a | n/a |  | * | n/a | n/a | 1/1 | 3/3 (100) |
| Rouse et al., 2024a | * | * | * | 3/3 |  | -- | 0/2 |  | * | * | n/a | 2/2 | 5/7 (71) |
| Rouse et al., 2024b | * | * | * | 3/3 |  | -- | 0/2 |  | * | * | n/a | 2/2 | 5/7 (71) |
| Sablik et al., 2025 | * | * | * | 3/3 |  | *- | 1/2 |  | * | * | * | 3/3 | 7/8 (88) |
| Samudra et al., 2020 | * | n/a | * | 2/2 |  | n/a | n/a |  | * | n/a | n/a | 1/1 | 3/3 (100) |
| Saykin et al., 1995 | * | n/a | * | 2/2 |  | n/a | n/a |  | * | - | * | 2/3 | 4/5 (80) |
| Schwarz et al., 2009 | * | n/a | * | 2/2 |  | n/a | n/a |  | * | * | n/a | 2/2 | 4/4 (100) |
| Seidenberg et al., 2002 | * | * | * | 3/3 |  | -- | 0/2 |  | * | n/a | n/a | 1/1 | 4/6 (67) |
| Sheldon et al., 2011 | * | * | * | 3/3 |  | ** | 2/2 |  | * | - | n/a | 1/2 | 6/7 (86) |
| Sroubek et al., 2025 | * | n/a | * | 2/2 |  | n/a | n/a |  | * | * | * | 3/3 | 5/5 (100) |
| St Laurent et al., 2009 | * | * | * | 3/3 |  | ** | 2/2 |  | * | - | n/a | 1/2 | 6/7 (86) |
| Struck et al., 2024 | * | * | * | 3/3 |  | -- | 0/2 |  | * | n/a | n/a | 1/1 | 4/6 (67) |
| Strutt et al., 2011 | - | n/a | * | 1/2 |  | n/a | n/a |  | * | n/a | n/a | 1/1 | 2/3 (67) |
| Tan et al., 2019 | * | * | * | 3/3 |  | *- | 1/2 |  | * | n/a | n/a | 1/1 | 5/6 (83) |
| Tavakol et al., 2024 | * | * | * | 3/3 |  | *- | 1/2 |  | * | n/a | n/a | 1/1 | 5/6 (83) |
| Tedrus et al., 2020 | * | * | * | 3/3 |  | -- | 0/2 |  | * | n/a | n/a | 1/1 | 4/6 (67) |
| Thivard et al., 2005 | * | * | * | 3/3 |  | -- | 0/2 |  | * | n/a | n/a | 1/1 | 4/6 (67) |
| Toledano et al., 2013 | * | * | * | 3/3 |  | ** | 2/2 |  | * | n/a | n/a | 1/1 | 6/6 (100) |
| Tracey et al., 2007 | * | n/a | * | 2/2 |  | n/a | n/a |  | * | n/a | n/a | 1/1 | 3/3 (100) |
| Trebuchon-Da Fonseca et al., 2009 | * | n/a | * | 2/2 |  | n/a | n/a |  | * | n/a | n/a | 1/1 | 3/3 (100) |
| Tröster et al., 1995 | * | -- | * | 2/3 |  | ** | 2/2 |  | * | n/a | n/a | 1/1 | 5/6 (83) |
| Trujillo-Pozo et al., 2013 | * | n/a | * | 2/2 |  | n/a | n/a |  | * | * | n/a | 2/2 | 4/4 (100) |
| Tudesco et al., 2010 | * | * | * | 3/3 |  | ** | 2/2 |  | * | n/a | n/a | 1/1 | 6/6 (100) |
| Vanli Yavuz et al., 2016 | * | n/a | * | 2/2 |  | n/a | n/a |  | * | - | - | 1/3 | 3/5 (60) |
| Vascouto et al., 2020 | * | n/a | * | 2/2 |  | n/a | n/a |  | * | n/a | n/a | 1/1 | 3/3 (100) |
| Viskontas et al., 2000 | * | * | * | 3/3 |  | ** | 2/2 |  | * | * | n/a | 2/2 | 7/7 (100) |
| Viskontas et al., 2002 | * | * | * | 3/3 |  | ** | 2/2 |  | * | * | n/a | 2/2 | 7/7 (100) |
| Visser et al., 2018 | * | * | * | 3/3 |  | ** | 2/2 |  | * | * | n/a | 2/2 | 7/7 (100) |
| Voets et al., 2006 | * | * | * | 3/3 |  | -- | 0/2 |  | * | n/a | n/a | 1/1 | 4/6 (67) |
| Vogt et al., 2018 | * | n/a | * | 2/2 |  | n/a | n/a |  | * | * | * | 3/3 | 5/5 (100) |
| Volfart et al., 2020 | * | * | * | 3/3 |  | ** | 2/2 |  | * | n/a | n/a | 1/1 | 6/6 (100) |
| Volfart et al., 2019 | * | * | * | 3/3 |  | ** | 2/2 |  | * | n/a | n/a | 1/1 | 6/6 (100) |
| Voltzenlogel et al., 2006 | * | * | * | 3/3 |  | ** | 2/2 |  | * | n/a | n/a | 1/1 | 6/6 (100) |
| Voltzenlogel et al., 2014 | * | * | * | 3/3 |  | ** | 2/2 |  | * | n/a | n/a | 1/1 | 6/6 (100) |
| Voltzenlogel et al., 2015 | * | * | * | 3/3 |  | ** | 2/2 |  | * | n/a | n/a | 1/1 | 6/6 (100) |
| Wang et al., 2010 | * | * | * | 3/3 |  | ** | 2/2 |  | * | n/a | n/a | 1/1 | 6/6 (100) |
| Wang et al., 2011 | * | n/a | * | 2/2 |  | n/a | n/a |  | * | n/a | n/a | 1/1 | 3/3 (100) |
| Wang et al., 2024a | * | * | * | 3/3 |  | ** | 2/2 |  | * | n/a | n/a | 1/1 | 6/6 (100) |
| Wang et al., 2024b | * | * | * | 3/3 |  | ** | 2/2 |  | * | n/a | n/a | 1/1 | 6/6 (100) |
| Waseem et al., 2015 | - | n/a | * | 1/2 |  | n/a | n/a |  | * | * | * | 3/3 | 4/5 (80) |
| Whiteside et al., 2010 | * | n/a | * | 2/2 |  | n/a | n/a |  | * | * | n/a | 2/2 | 4/4 (100) |
| Whitten et al., 2021 | * | n/a | * | 2/2 |  | n/a | n/a |  | * | n/a | n/a | 1/1 | 3/3 (100) |
| Williams Roberson et al., 2020 | * | n/a | * | 2/2 |  | n/a | n/a |  | * | n/a | n/a | 1/1 | 3/3 (100) |
| Yang et al., 2016 | * | n/a | * | 2/2 |  | n/a | n/a |  | * | * | * | 3/3 | 5/5 (100) |
| Yucus et al., 2007 | * | - | * | 2/3 |  | -- | 0/2 |  | * | * | n/a | 2/2 | 4/7 (57) |
| Zalonis et al., 2017 | * | n/a | * | 2/2 |  | n/a | n/a |  | * | n/a | n/a | 1/1 | 3/3 (100) |
| Zannino et al., 2020 | * | n/a | * | 2/2 |  | n/a | n/a |  | * | * | n/a | 2/2 | 4/4 (100) |

^a^Items not relevant for specific studies are denoted by n/a. This includes studies without a healthy control group or a longitudinal component.
